# Tumor *NLRP3* Amplification Promotes Immunotherapy Resistance by Suppressing MHC Class I Expression

**DOI:** 10.1101/2025.10.06.25337175

**Authors:** Bala Theivanthiran, Nagendra Yarla, Kaylee Villareal, Y-Van Nguyen, Linda Cao, Ernesto P. Calderin, Michael P. Plebanek, Kyra Majors, Emily Bolch, Douglas B. Johnson, Hope Uronis, John H. Strickler, Nicholas C. DeVito, Brent A. Hanks

## Abstract

Immunotherapy resistance remains a major challenge in immuno-oncology. We have previously demonstrated that a tumor-intrinsic NLRP3 inflammasome signaling pathway promotes adaptive immunotherapy resistance by inducing the recruitment of granulocytic myeloid-derived suppressor cells. We now confirm that elevated tumor NLRP3 signaling activity correlates with checkpoint inhibitor resistance in several independent cohorts of stage III and IV melanoma patients, as well as in advanced gastroesophageal (GE) adenocarcinoma patients. *In situ* hybridization studies further demonstrate that tumor *NLRP3* copy-number gain is observed in immunotherapy resistant melanomas and GE adenocarcinomas harboring enhanced NLRP3 signaling activity. Spatial transcriptomic analysis of GE adenocarcinoma tissues reveals that NLRP3 signaling activity inversely correlates with NLRC5 and MHC class I-associated gene expression. Indeed, pre-clinical models of melanoma and GE adenocarcinoma demonstrate that *Nlrp3* amplification suppresses NLRC5-mediated MHC class I upregulation, while pharmacologic NLRP3 inhibition augments tumor MHC class I surface levels. Upon activation, tandem mass spectrometry-supported signaling studies reveal that NLRP3 binds to and inhibits STAT1 dimerization, nuclear translocation, and NLRC5 transcription. Consistent with these findings, inhibiting the NLRP3 inflammasome augments tumor STAT1-NLRC5 signaling and overcomes anti-PD-1 resistance in an orthotopic model of chromosomal instability (CIN) gastric adenocarcinoma. This work indicates that the tumor NLRP3 inflammasome signaling pathway merits further clinical study as a therapeutic target and a source of companion biomarkers for overcoming checkpoint inhibitor resistance in cancer patients.

## Introduction

While the introduction of checkpoint inhibitor immunotherapies targeting PD-1 have revolutionized clinical oncology, this benefit has been limited to less than 20% of cancer patients ^1^. In addition, multiple combination regimens including anti-PD-1 immunotherapy have either failed to improve or only modestly improve outcomes over anti-PD-1 monotherapy for a variety of indications ^2,3^. The identification of novel immunotherapeutic targets and companion predictive biomarkers allowing for rational selection of tailored combination regimens for the individual patient is now a priority for advancing the field of immuno-oncology ^4^.

Using an array of pre-clinical tumor models, we have previously demonstrated that activation of CD8^+^ T cells in response to PD-1 blockade stimulates a tumor-intrinsic NOD-, LRR-, and pyrin-domain-containing protein 3 (NLRP3) inflammasome-heat shock protein-70 (HSP70) signaling axis that generates a CXCR2-dependent chemokine gradient, facilitating the recruitment of granulocytic myeloid-derived suppressor cells (PMN-MDSCs) into the tumor bed as well as their accumulation in more distant tissues ^5,6^. Both genetic silencing and pharmacologic inhibition of either tumor-expressed NLRP3 or HSP70 enhances CD8^+^ T cell tumor infiltration and augments responses to anti-PD-1 immunotherapy in an autochthonous model of melanoma. We have further shown that elevated baseline levels of activation of the tumor-intrinsic NLRP3 inflammasome correlates with resistance to anti-PD-1 immunotherapy in a cohort of stage IV melanoma patients ^6^. These studies also revealed that the cytoplasmic domain of PD-L1 can trigger tumor NLRP3 inflammasome activation via a STAT3-dsRNA-dependent protein kinase R (PKR) signaling axis, thus linking tumor PD-L1 expression with NLRP3-dependent adaptive resistance to anti-PD-1 immunotherapy ^5^. Indeed, genetic silencing of tumor PD-L1 eliminates the activation of the NLRP3-HSP70 pathway, suggesting that the tumor-intrinsic NLRP3-HSP70 signaling pathway may contribute to anti-PD-1 resistance in select PD-L1-positive tumors ^5^. Interestingly, *NLRP3* copy-number gains have been observed in an array of solid tumors ^7^. However, it is currently unclear if the genetic state of *NLRP3* influences carcinogenesis or determines whether tumor-intrinsic NLRP3 activation occurs in response to anti-PD-1 immunotherapy.

Previous studies have shown that NOD-, LRR-, and CARD-containing 5 (NLRC5) regulates various genes that play a role in MHC class I transcription ^8^. As such, tumor expression of NLRC5 has been positively correlated with tumor MHC class I expression, tumor-infiltrating CD8^+^ T cell activation levels, and the overall survival of several populations of cancer patients ^9^. Functional loss of various genes associated with MHC class I antigen processing and presentation by various tumor types often serves as a mechanism for evading anti-tumor immunity, including responses to checkpoint inhibitor immunotherapy ^10–12^. Unfortunately, our ability to overcome this evasion strategy to enhance anti-tumor immunity has been limited ^13–15^.

Unlike in melanoma, there have been modest benefits achieved with anti-PD-1 immunotherapy in advanced gastroesophageal (GE) adenocarcinoma patients and these responses are rarely durable, leaving patients with few compelling treatment options beyond first line therapy ^16^. Even with available combination immunotherapy regimens, the median overall survival of advanced GE adenocarcinoma patients remains limited to approximately 14 months and the incidence of this cancer is now significantly increasing, particularly in younger patient populations ^17–19^.

Here, we explore the role of *NLRP3* genetic amplification in driving tumor-mediated immune escape during anti-PD-1 immunotherapy in melanoma as well as GE adenocarcinoma patients and demonstrate tumor-intrinsic NLRP3 activation to suppress NLRC5-mediated transcription of the MHC class I antigen presentation machinery of these tumor types. Pharmacological inhibition of NLRP3 is shown to reverse MHC class I downregulation and overcome resistance to anti-PD-1 immunotherapy in autochthonous and orthotopic models of these malignancies, respectively. These studies provide additional data supporting the exploration of the tumor-intrinsic NLRP3 inflammasome pathway as a promising source of novel therapeutic targets and companion biomarkers to overcome anti-PD-1 resistance in difficult to treat patient populations.

## Results

### Tumor-intrinsic NLRP3 Inflammasome Activity and Tumor NLRP3 Amplification is Associated with Anti-PD-1 Resistance in Advanced Melanoma Patients

Using a NLRP3-ASC proximity ligation assay (PLA) as a surrogate marker for NLRP3 inflammasome activity, we have previously shown that elevated baseline tumor-intrinsic NLRP3 signaling activity correlates with resistance to checkpoint inhibitor immunotherapy in naïve stage IV melanoma patients ^6^. In order to verify these findings, we examined baseline tumor tissue specimens harvested from an independent cohort of stage IV melanoma patients treated at an outside institution using the same NLRP3-ASC PLA (**Fig. 1a, Supplementary Table 1**). This work verified our previous findings, demonstrating that elevated baseline tumor NLRP3 signaling activity correlates with resistance in patients that have received either anti-PD-1 monotherapy or combination anti-PD-1/anti-CTLA-4 therapy. Indeed, our cumulative data shows that elevated baseline tumor NLRP3 signaling activity correlates with diminished progression-free survival (PFS) in stage IV melanoma patients undergoing checkpoint inhibitor immunotherapy (**Fig. 1b, Supplementary Table 1**).

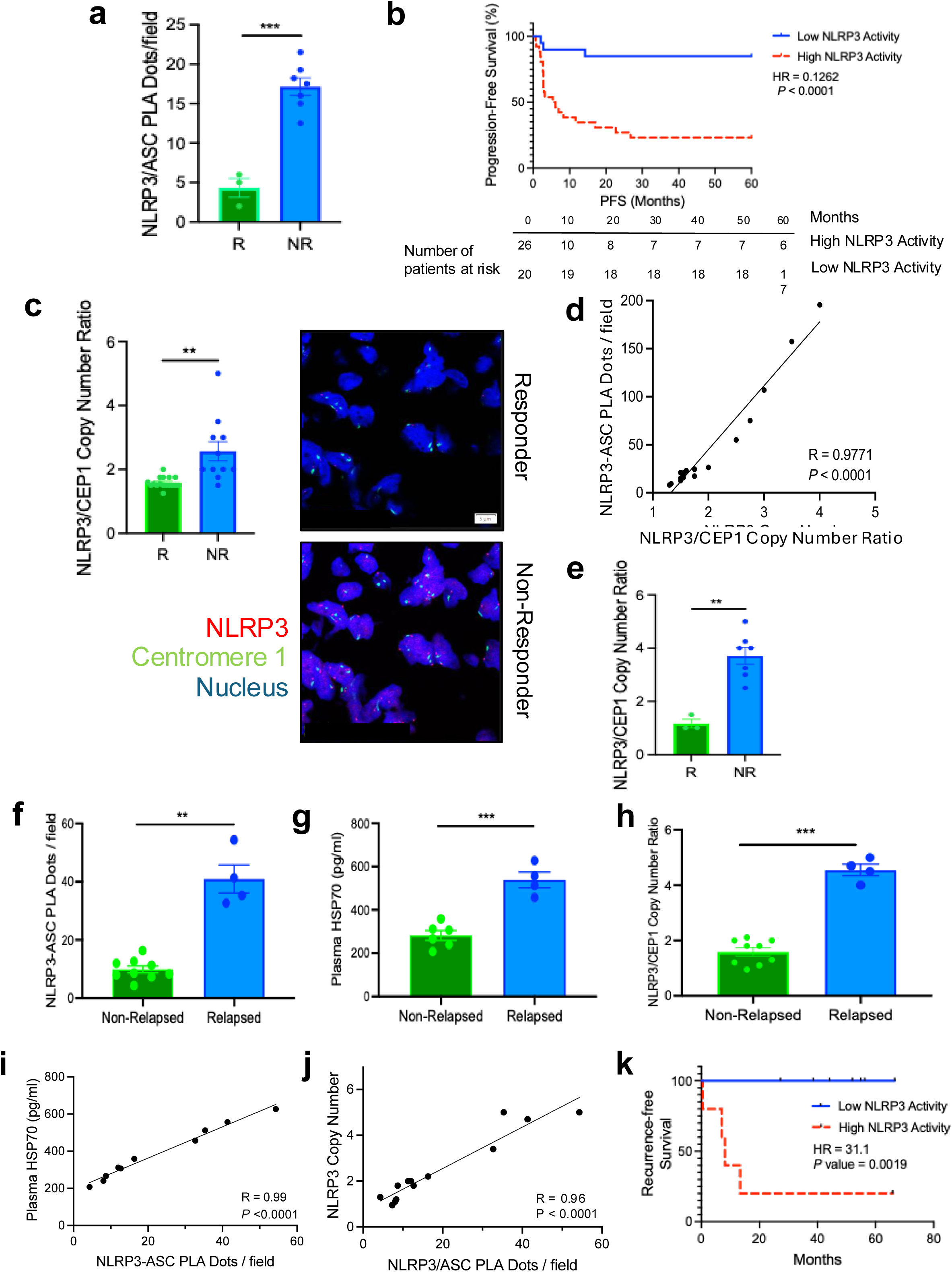

We reasoned that an underlying genetic mechanism was likely determining whether a tumor could leverage the NLRP3 inflammasome pathway to evade anti-tumor immunity. Prior studies have noted that *NLRP3* genetic amplification occurs in several solid tumors ^7^. Indeed, ∼16% of cutaneous melanomas, including both early- and late-stage disease, in The Cancer Genome Atlas (TCGA) harbor amplified *NLRP3* ^6,20^. Based on these data, we performed a *NLRP3* fluorescence *in situ* hybridization (FISH) assay on baseline tumor tissue specimens derived from stage IV melanoma patients initiating either anti-PD-1 monotherapy or combination anti-PD-1/anti-CTLA-4. This analysis showed copy number gains of *NLRP3* associated with resistance to first-line checkpoint inhibitor immunotherapy (**Fig. 1c, Supplementary Table 1**). Indeed, *NLRP3* gene copy number was found to correlate closely with NLRP3 signaling activity in melanoma tissues, suggesting that copy number gain is the primary genetic mechanism influencing tumor NLRP3 signaling activity (**Fig. 1d**). We further verified these findings by performing *NLRP3* FISH assays on baseline tissue specimens derived from an independent cohort of stage IV melanoma patients managed with checkpoint inhibitor immunotherapy (**Fig. 1e**). Overall, this work suggests that somatic copy number gains of *NLRP3* by melanoma tissues influences tumor NLRP3 signaling activity and promotes resistance to checkpoint inhibitor immunotherapy.

There is currently a significant need for the development of predictive biomarkers for managing stage III melanoma patients, as many of these patients are exposed to checkpoint inhibitor immunotherapy placing the patient at risk for developing significant immune-related adverse events while providing no clinical benefit. As a result, we conducted NLRP3-ASC PLA and NLRP3 FISH assays on baseline tumor specimens along with an HSP70 enzyme-linked immunosorbent assay (ELISA) on baseline plasma specimens derived from stage III melanoma patients prior to initiating adjuvant anti-PD-1 immunotherapy. These studies demonstrated elevated tumor NLRP3 signaling activity, increased baseline plasma levels of HSP70, and tumor *NLRP3* amplification to be associated with anti-PD-1 failure and melanoma recurrence (**Figs. 1f-h, Supplementary Table 2**). Again, we found baseline plasma HSP70 levels to correlate closely with tumor NLRP3 activity levels and, in turn, with *NLRP3* amplification levels in this cohort of stage III melanoma patients (**Figs. 1i, j**). Consistent with these findings, stage III melanoma patients with tumor NLRP3 activity levels above the mean are associated with inferior recurrence-free survival relative to those harboring melanomas with NLRP3 activity below the mean (**Fig. 1k**). These cumulative data are consistent with our previously reported studies and suggest that tumor-intrinsic NLRP3 signaling activity supports immune evasion and promotes checkpoint inhibitor immunotherapy resistance in melanoma ^5,6^.

### A NLRP3 Transcriptional Signature Correlates with Resistance to Anti-PD-1 Immunotherapy in Advanced Melanoma Patients

In order to examine the relationship between activation of the tumor-intrinsic NLRP3 inflammasome and immunotherapy resistance in publicly available clinical datasets, we constructed an unbiased transcriptional signature of NLRP3 activation in melanoma tissues by utilizing a genetically engineered murine model of melanoma (**Fig. 2a**). This included capturing upregulated genes in BRAF^V600E^PTEN^-/-^ melanomas modified via CRISPRa to exhibit enhanced *Nlrp3* transcriptional activity (BRAF^V600E^PTEN^-/-^NLRP3a) and integrating this gene set with the downregulated genes in a transgenic Tyr-cre;BRAF^V600E^PTEN^-/-^NLRP3^-/-^ tumor model exhibiting melanoma-restricted ablation of *Nlrp3* (**Extended Data Fig. 1**). Based on this approach, we generated a 48-gene transcriptional profile reflecting tumor-intrinsic NLRP3 activation and found this ‘NLRP3-tsig’ signature to positively correlate with a PMN-MDSC transcriptional signature in melanoma tissues using single sample gene set enrichment analysis (ssGSEA) (**Fig. 2b, Supplementary Table 3**). We then performed receiver-operator characteristic (ROC) curve analyses of NLRP3-tsig applied to independent previously reported datasets of stage IV melanoma patients treated with anti-PD-1 immunotherapy ^21^. This work showed the NLRP3 signature to predict for resistance to anti-PD-1 immunotherapy with an area under the curve (AUC) value of 0.724 in the Hugo et al dataset (26 patients) and an AUC value of 0.679 in the Gide et al dataset (29 patients) (**Fig. 2c**) ^21,22^. Indeed, the NLRP3-tsig signature outperformed both the PMN-MDSC signature and a M-MDSC signature in predicting response to anti-PD-1 immunotherapy in melanoma patients (**Fig. 2d)** ^23,24^. Importantly, the NLRP3-tsig signature also performed favorably to previously reported signatures in predicting anti-PD-1 resistance in advanced melanoma, including the ICBresistance, X18signature, TIDE, IMPRES, and IPRES signatures in the Hugo et al. dataset, as well as the larger Lee et al. dataset (43 patients) (**Figs. 2e, f**) ^21,25–29^. We further found this approach to generate a NLRP3-associated transcriptional signature that is superior in predicting resistance relative to a previously developed NLRP3pancancersig signature constructed from various components of the NLRP3 signaling pathway in both of these datasets (**Fig. 2e, f**) ^30^. Finally, our studies indicate the NLRP3-tsig signature also correlates with recurrence-free survival in the TCGA melanoma dataset (**Fig. 2g**). Our review of the TCGA has revealed that several other solid tumor malignancies also exhibit *NLRP3* amplification ^31^. Based on these cumulative findings, we next sought to verify whether tumor-intrinsic NLRP3 signaling activity may also contribute to immunotherapy resistance in a more refractory population of advanced GE adenocarcinoma patients.

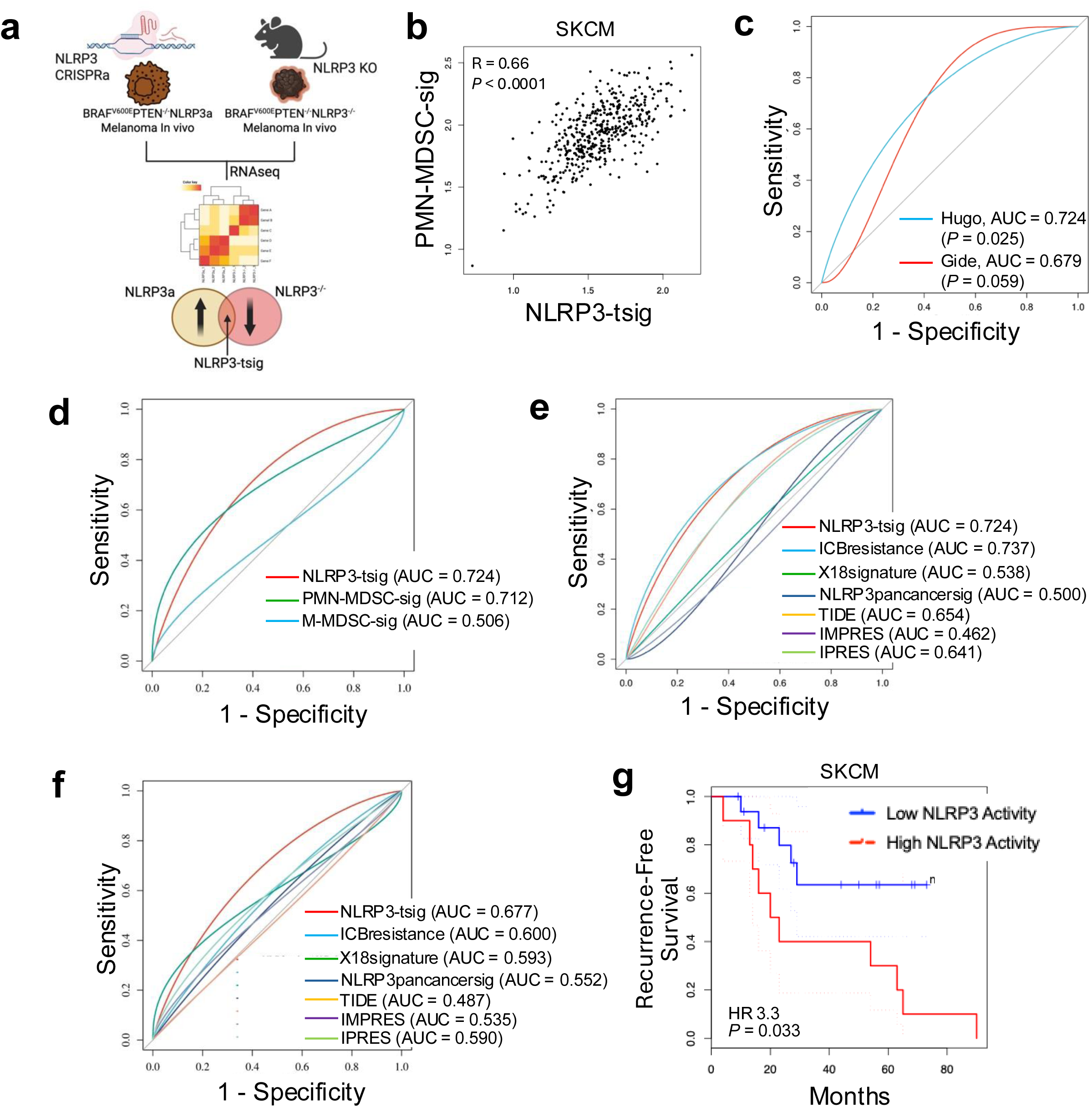

### Tumor-intrinsic NLRP3 Inflammasome Activity and Tumor NLRP3 Amplification is Associated with ChemoImmunotherapy Resistance in Advanced Gastroesophageal Cancer Patients

The phase II KeyLARGO clinical trial evaluated first-line capecitabine/oxaliplatin (CAPOX) chemotherapy with pembrolizumab (anti-PD-1 antibody) in unresectable stage III or stage IV microsatellite-stable (MSS), HER2-negative GE adenocarcinoma patients following an 8-day lead-in of pembrolizumab monotherapy ^32^. In this study, both tissue and blood specimens were acquired at baseline and at day 8 prior to initiating chemotherapy, providing a window of opportunity to analyze the impact of anti-PD-1 immunotherapy alone on GE adenocarcinoma tissues (**Figs. 3a, b, Supplementary Table 4**). Notably, prior studies observed no relationship between PD-L1 combined positive score (CPS) testing in these tumor tissue specimens and treatment response in this patient cohort (**Extended Data Fig. 2a**) ^32^. Paired tissue and blood specimens were harvested from 10 patients, evaluated by a board-certified gastrointestinal pathologist, and analyzed utilizing the NLRP3-ASC PLA and the HSP70 ELISA, respectively. Baseline specimens were also evaluated by *NLRP3* FISH. Based on immunofluorescence microscopy, we found an increased number of tumor-infiltrating CD8^+^ T cells at day 8 relative to baseline following treatment with pembrolizumab (**Fig. 3c**). This analysis also demonstrated an increased number of baseline tumor-infiltrating CD8^+^ T cells in responders versus non-responders (**Fig. 3d**). Furthermore, this increase in tumor-infiltrating CD8^+^ T cells following pembrolizumab immunotherapy was only noted to occur in responding patients (**Fig. 3e**). Using elastase as a marker of neutrophils or polymorphonuclear cells (PMNs), an increase in this cell population was also noted to occur by day 8 following pembrolizumab immunotherapy (**Fig. 3f**). Baseline PMNs were elevated in non-responders relative to responding patients and an increase in PMNs with pembrolizumab immunotherapy was only noted to occur in non-responding patients (**Figs. 3g, h**). Assuming that these PMNs are functionally suppressive, and therefore consistent with PMN-MDSCs, these data are compatible with our prior work in melanoma showing an increase in tumor-infiltrating PMN-MDSCs upon treatment with anti-PD-1 antibody in resistant tumors ^5^. As we previously showed that this effect was secondary to activation of the tumor-intrinsic NLRP3 inflammasome, we assessed these tissue specimens using the NLRP3-ASC PLA and found the tumors of non-responders to exhibit elevated NLRP3 signaling activity (**Fig. 3i**). In light of this relationship, we then evaluated the accompanying plasma samples from an expanded patient cohort derived from this study and found elevated baseline HSP70 plasma levels to correlate with progressive disease (**Fig. 3j**). Similar to our previous findings related to PMNs, we also found only non-responding patients to demonstrate an increase in plasma HSP70 upon treatment with pembrolizumab immunotherapy (**Fig. 3k**). Notably, baseline levels of CD8^+^ T cells and PMNs, as well as plasma HSP70, correlated with tumor NLRP3 signaling activity based on the NLRP3-ASC PLA (**Extended Data Fig. 2b**). Given our prior data implicating *NLRP3* amplification as an important factor determining response to checkpoint inhibitor immunotherapy in melanoma (**Figs. 1c-e**), we also performed *NLRP3* FISH analysis of these GE adenocarcinoma tissue specimens. Consistent with our previous studies, this work also determined that GE adenocarcinomas exhibiting baseline *NLRP3* amplification did not respond to this chemoimmunotherapy regimen (**Fig. 3l**). Indeed, patients harboring GE adenocarcinomas exhibiting elevated levels of *NLRP3* amplification were found to be associated with an inferior PFS (HR = 0.261, *P* = 0.0087) and overall survival (OS, HR = 0.253, *P* = 0.0091) relative to those patients with non-amplified tumors (**Fig. 3m**). We then utilized the NLRP3-tsig signature described above and found it to also correlate with the PMN-MDSC signature and with recurrence-free survival in the gastric adenocarcinoma TCGA dataset (**Figs. 3n,o**). Although these data are derived from a limited patient cohort, the results are consistent with our prior melanoma studies, suggesting that tumor *NLRP3* amplification and enhanced NLRP3 signaling correlate with anti-PD-1 resistance in GE adenocarcinoma.

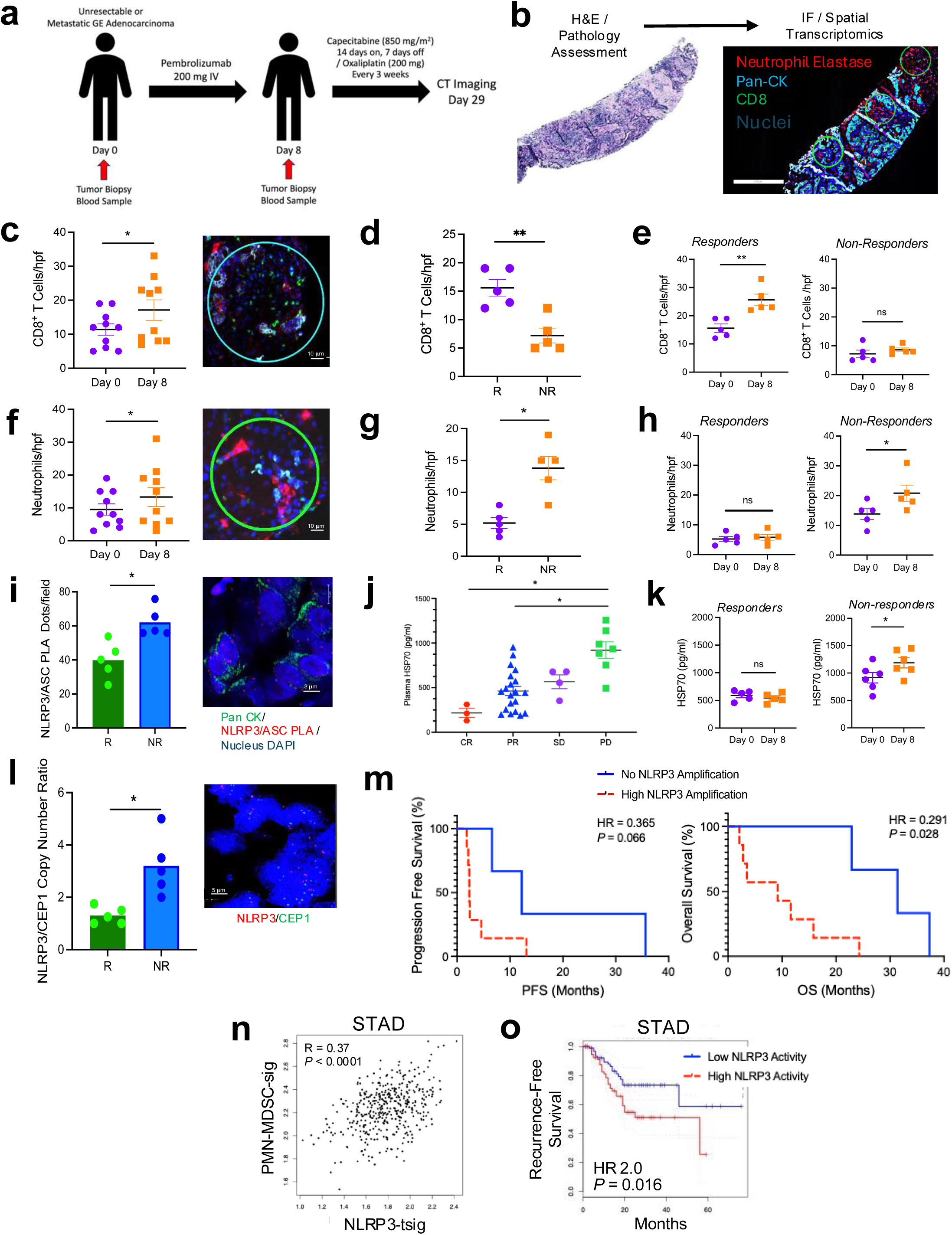

### Tumor-intrinsic NLRP3 Expression Levels Inversely Correlate with NLRC5 and MHC Class I Expression Levels

Applying spatial transcriptomics to day 0 and day 8 paired tumor specimens derived from the GE adenocarcinoma patients undergoing anti-PD-1 immunotherapy, we found *NLRP3* expression levels to be elevated in non-responding relative to responding patients (**Fig. 4a**). We further found increased expression of genes associated with the NLRP3 inflammasome complex in baseline tumor tissue specimens based on KEGG pathway analysis in non-responding patients while these tumor tissues also demonstrated increased NLRP3 inflammasome expression levels by day 8 of anti-PD-1 therapy relative to day 0 (**Fig. 4b**). Other transcripts associated with the tumor-intrinsic NLRP3 signaling pathway, such as *CXCL5*, were also found to be upregulated in non-responding patients (**Extended Data Fig. 3a**) ^5^. This transcriptional analysis further revealed several genes associated with MHC class I antigen processing and presentation to be downregulated by day 8 of anti-PD-1 therapy, including *B2M* (β2-microglobulin), *TAP1/2* (transporter-associated with antigen processing-1/2), and several *HLA* (human leukocyte antigen) alleles (**Fig. 4c**). Interestingly, a decrease in *HLA-A* and *HLA-B* expression by day 8 of anti-PD-1 immunotherapy was also found to be restricted to the tumor tissues harvested from non-responders, while this was not observed in the tumor tissues of responders (**Fig. 4d, *top***). Indeed, these observations were concordant with pan-MHCI immunohistochemistry (IHC) studies, where increased expression levels of MHCI were noted exclusively in the tumor tissues of responders (**Fig. 4d, *bottom***). As we observed consistent changes across several MHC class I-associated genes, we also investigated the MHC class I transcriptional transactivator, NOD-like receptor family CARD domain containing-5 (NLRC5). These studies confirmed NLRC5 expression levels to also be diminished in day 8 relative to day 0 tumor tissues while IHC showed elevated NLRC5 expression levels in responding relative to non-responding GE adenocarcinoma patients (**Figs. 4e, f**). Interestingly, transcripts associated with cellular stress-related pathways were also found to be increased in the same non-responding patients following anti-PD-1 immunotherapy (**Fig. 4g**) ^33^.

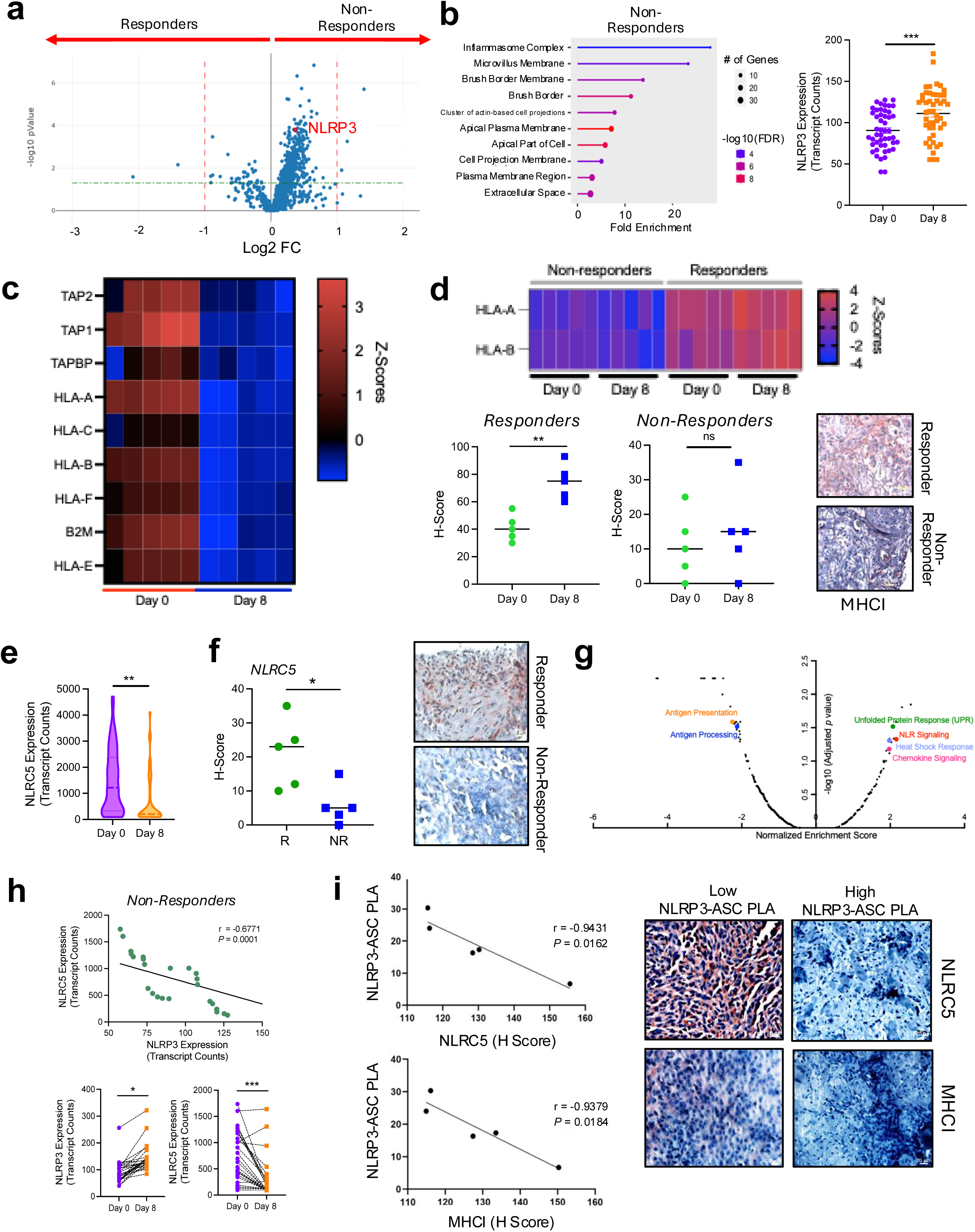

Given our prior studies revealing elevated NLRP3 expression levels in day 8 tumor tissues, we hypothesized that tumor-intrinsic NLRP3 signaling may suppress NLRC5 expression. Indeed, this relationship was further supported by findings showing that tumor tissues with elevated *NLRP3* expression also exhibit lower *NLRC5* expression levels (**Fig. 4h, *top***). While *NLRP3* expression levels uniformly increased in non-responders in day 8 tumor tissues, we found *NLRC5* expression levels to decrease in these same tumors by day 8 (**Fig. 4h, *bottom***). Further analysis of individual tumor regions with increased CD8^+^ T cells and PMNs also showed a trend toward decreased *NLRC5* and *STAT1* expression relative to those regions harboring low levels of PMNs (**Extended Data Fig. 3b**). To confirm that this relationship between NLRP3 and NLRC5/MHCI also existed in other tumors beyond GE adenocarcinoma, we turned again to tumor tissue specimens harvested from melanoma patients and performed the NLRP3-ASC PLA described above along with NLRC5 and pan-MHCI IHC. This study verified that the tumor-intrinsic NLRP3 inflammasome activity inversely correlates with both NLRC5 and MHCI expression, suggesting that NLRP3 signaling activity may also suppress anti-tumor immunity by downregulating MHC class I antigen processing and presentation (**Fig. 4i**).

We have previously demonstrated that the tumor-intrinsic NLRP3 inflammasome pathway triggers the recruitment of PMN-MDSCs into the TME and others have reported a link between tumor-infiltrating PMN-MDSCs and MHC class I downregulation ^34^. However, we found no effect of PMN-MDSC ablation on tumor MHC class I expression levels in the transgenic BRAF^V600E^PTEN^-/-^ melanoma model, suggesting that NLRP3 inflammasome signaling may directly downregulate NLRC5/MHC class I expression by tumors (**Extended Data Fig. 3c**).

### Tumor-intrinsic NLRP3 Signaling Activity Suppresses NLRC5 and MHC Class I Expression Levels in Tumors

In order to investigate whether NLRP3 signaling suppresses NLRC5, and therefore MHC class I gene expression in tumor tissues, we used a previously engineered murine BRAF^V600E^PTEN^-/-^ melanoma cell line harboring transcriptionally amplified *Nlrp3* (BRAF^V600E^PTEN^-/-^NLRP3a) ^6^. RNAseq transcriptional analysis of BRAF^V600E^PTEN^-/-^ NLRP3a melanoma demonstrated a downregulation in the expression of genes associated with antigen processing and presentation relative to control BRAF^V600E^PTEN^-/-^-NTC tumors (**Fig. 5a**). While Western blot studies showed IFN-γ stimulation to enhance NLRC5 expression levels in a control BRAF^V600E^PTEN^-/-^ melanoma cell line (BRAF^V600E^PTEN^-/-^NTC), this increase was suppressed in the BRAF^V600E^PTEN^-/-^ NLRP3a melanoma cell line (**Fig. 5b**). Consistent with these findings, we also found that a small molecule inhibitor of the NLRP3 inflammasome, MCC950, enhanced NLRC5 expression levels in both murine and human melanoma cell lines, as well as NLRC5 in the KPCY murine pancreatic ductal adenocarcinoma cell line (**Fig. 5c, Extended Data Figs. 4a, b**) ^35^. To determine whether corresponding alterations in MHC class I surface expression are observed in response to NLRP3 signaling activity, we performed pan-MHCI flow cytometry analysis of BRAF^V600E^PTEN^-/-^NLRP3a and BRAF^V600E^PTEN^-/-^NTC melanomas following their surgical resection and observed a significant diminishment in MHC class I surface expression in *Nlrp3*-amplified melanomas (**Fig. 5d, *left***). Indeed, pharmacologic inhibition of NLRP3 of BRAF^V600E^PTEN^-/-^ melanomas *in vivo* and the KPCY pancreatic adenocarcinoma cell line *in vitro* correspondingly resulted in enhanced levels of MHC class I surface expression (**Fig. 5d, *right*, Extended Data Fig. 4c**).

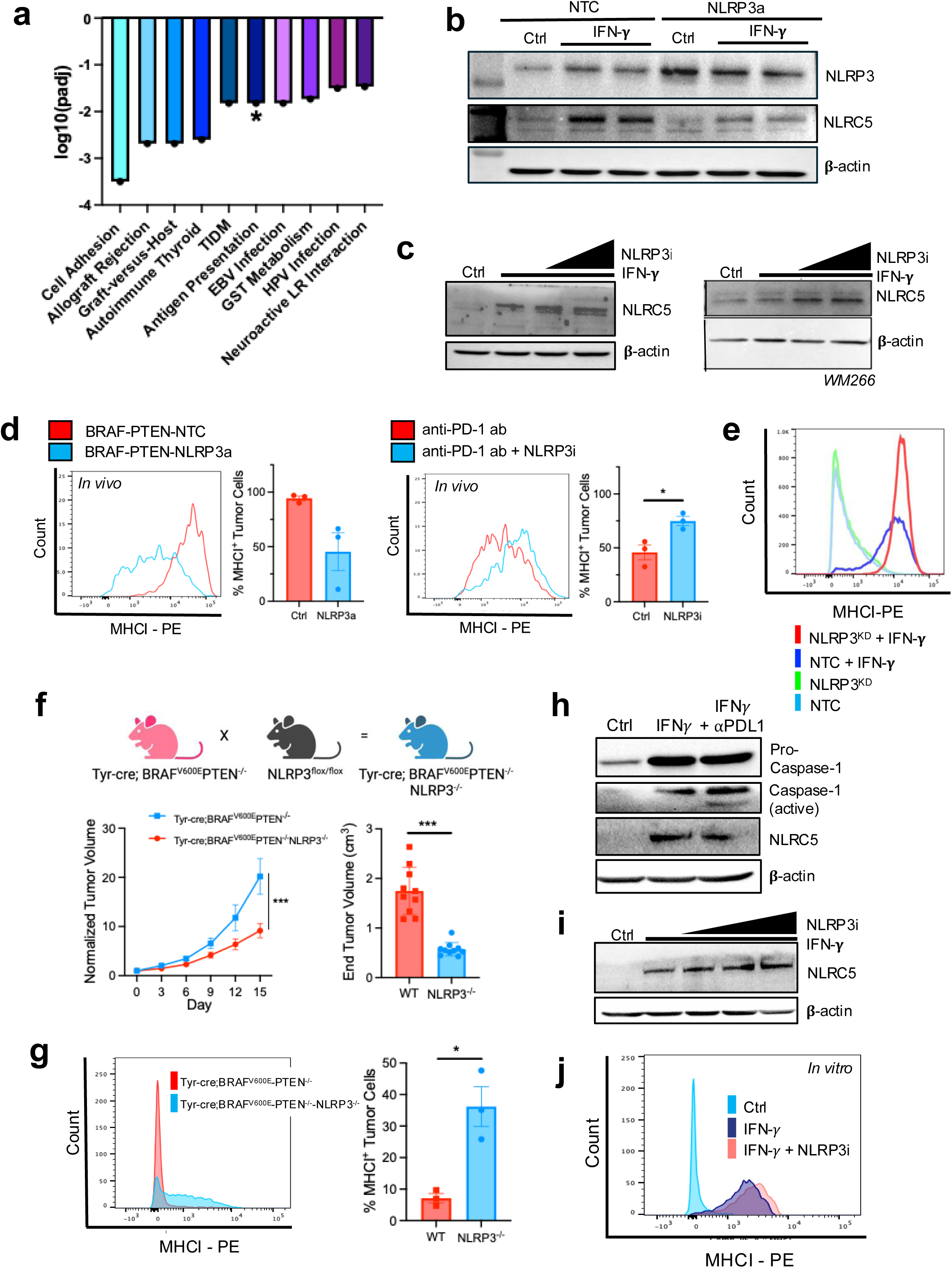

We have previously shown that PD-L1 triggers activation of the NLRP3 inflammasome signaling pathway in tumors ^5^. As a result, we performed Western blot studies in *Cd274*-silenced BRAF^V600E^PTEN^-/-^ melanoma cell lines (BRAF^V600E^PTEN^-/-^ PDL1^KD^) and noted an increase in NLRC5 expression levels, a finding that was phenocopied by a *Nlrp3*-silenced BRAF^V600E^PTEN^-/-^ melanoma cell line (**Extended Data Fig. 4d**). These findings were consistent with an observed increase in surface MHC class I in response to IFN-γ stimulation in both BRAF^V600E^PTEN^-/-^NLRP3^KD^ and BRAF^V600E^PTEN^-/-^PDL1^KD^ tumor cell lines (**Fig. 5e, Extended Data Fig. 4e**). In order to examine this pathway more rigorously *in vivo*, we crossed Tyr-cre;BRAF^V600E^PTEN^-/-^ mice with NLRP3^flox/flox^ mice to generate an autochthonous melanoma model silenced for melanocyte-specific *Nlrp3* expression (Tyr-cre;BRAF^V600E^PTEN^-/-^NLRP3^-/-^) (**Fig. 5f, *top***). Melanoma tissue-restricted knockout of *Nlrp3* expression was first confirmed and diminished expression of genes known to play a role in PMN recruitment were also verified in BRAF^V600E^PTEN^-/-^NLRP3^-/-^ tumors (**Extended Data Figs. 1a, b**). In line with our previous data, we found BRAF^V600E^PTEN^-/-^NLRP3^-/-^ tumor growth to be suppressed relative to control BRAF^V600E^PTEN^-/-^ tumors (**Fig. 5f, *bottom***). This was further consistent with a significant increase in MHC class I surface expression in BRAF^V600E^PTEN^-/-^NLRP3^-/-^ tumors *in situ* compared with control BRAF^V600E^PTEN^-/-^ tumors (**Fig. 5g**). As evidence of this NLRP3-NLRC5 axis was originally detected in tissue specimens derived from GE adenocarcinoma patients, we conducted a series of studies in a murine model of chromosomal instability (CIN) gastric adenocarcinoma derived from a recently engineered Anxa10-CreER^T2^;Kras^G12D/+^;Tp53^R172H/+^;Smad4^f/f^ transgenic mouse model (KPS), which we also determined to be *Nlrp3* amplified with a Nlrp3/CEP1 ratio of 2 (**Extended Data Fig. 5a**) ^36^. Studies confirmed IFN-γ/anti-PD-L1 treatment to induce NLRP3 and caspase-1 activation and for this to be associated with enhanced HSP70 release and suppressed NLRC5 expression in the CIN KPS gastric cancer cell line *in vitro* (**Fig. 5h, Extended Data Figs. 5b, c**). We also verified anti-PD-1 immunotherapy to induce NLRP3 activation and HSP70 release in the orthotopic CIN KPS gastric cancer model, consistent with our observations in melanoma (**Extended Data Figs. 5d, e**). In line with these findings, we observed NLRP3 inhibition with MCC950 to increase levels of NLRC5 and MHC class I expression *in vitro* (**Figs. 5i, j**). Together, this work describes a previously unrecognized role for the NLRP3 inflammasome in suppressing NLRC5 and the MHC class I antigen processing and presentation machinery of tumors.

### Tumor-intrinsic NLRP3 Inflammasome Inhibits NLRC5 Expression by Inhibiting STAT1-mediated Transcription

We initially hypothesized that the tumor-intrinsic NLRP3 pathway may be eliciting the upregulation of a soluble factor that suppresses NLRC5-MHC class I tumor expression, prompting experiments demonstrating that the conditioned media of *Nlrp3*-amplified BRAF^V600E^PTEN^-/-^ melanoma cells modestly suppresses IFN-γ-dependent induction of NLRC5 and MHC class I expression (**Extended Data Figs. 6a, b**). Since soluble HSP70 is released downstream of tumor-intrinsic NLRP3 activation, we compared NLRC5-MHC class I expression between BRAF^V600E^PTEN^-/-^HSP70^-/-^ versus BRAF^V600E^PTEN^-/-^NTC control cells and found NLRC5 expression to be moderately elevated in HSP70-ablated tumor cells (**Extended Data Fig. 6c)**. Indeed, there was a slight increase in MHC class I surface levels in HSP70-silenced BRAF^V600E^PTEN^-/-^ melanoma cells, as well as BRAF^V600E^PTEN^-/-^ melanoma cells treated with an anti-HSP70 antagonistic antibody (**Extended Data Fig. 6d**). However, given the modest overall changes in tumor NLRC5-MHC class I expression observed, we initiated studies to explore other pathways that may be regulated more directly by NLRP3. Therefore, we immunoprecipitated NLRP3 from BRAF^V600E^PTEN^-/-^ melanoma cells following NLRP3 activation via IFN-γ/anti-PD-L1 ab treatment and analyzed these products by tandem mass spectrometry versus untreated BRAF^V600E^PTEN^-/-^ melanoma cells. Of the 27 unique proteins found to bind to NLRP3 upon IFN-γ/anti-PD-L1 ab activation in tumor cells, signal transducer and activator of transcription 1 (STAT1) was determined to be the most abundant (**Fig. 6a, Extended Data Fig. 7a, Supplementary Table 5**). Since NLRC5 is transcriptionally regulated by STAT1, we conducted a series of co-immunoprecipitation experiments to confirm NLRP3-STAT1 binding in the BRAF^V600E^PTEN^-/-^ melanoma cell line (**Fig. 6b**). We noted that IFN-γ/anti-PD-L1 ab treatment of BRAF^V600E^PTEN^-/-^ melanoma cells suppressed phosphorylated STAT1 levels, an effect that was enhanced with NLRP3-FLAG expression (**Fig. 6c**). Indeed, time course studies showed NLRP3-FLAG expression to more rapidly suppress both total and phosphorylated levels of STAT1 in BRAF^V600E^PTEN^-/-^ melanoma cells (**Fig. 6d**). Based on these findings, we looked for and found no evidence that NLRP3-STAT1 binding promotes STAT1 proteasomal degradation (**Extended Data Fig. 7b**). However, additional Western blot studies following nuclear-cytoplasmic fractionation showed NLRP3-FLAG expression to nearly eliminate total STAT1 levels in the nucleus (**Fig. 6e**). Based on these findings, we conducted native non-denaturing Western blots and found NLRP3 to suppress STAT1 dimerization, a process that is necessary for STAT1 nuclear translocation (**Fig. 6f**) ^37^. Consistent with these findings, we used a luciferase reporter system to further demonstrate that NLRP3 inhibits STAT1-dependent transcriptional activity (**Fig. 6g**). As STAT1 is known to auto-regulate its own transcription, we further confirmed a downregulation of STAT1 and other MHC class I transcript levels in both BRAF^V600E^PTEN^-/-^ melanoma cells and BRAF^V600E^PTEN^-/-^ melanoma tissues harboring amplified *Nlrp3* (**Fig. 6h**). Notably, we also determined NLRP3 activation to similarly suppress STAT1 phosphorylation and *Nlrc5* expression in primary bone marrow-derived dendritic cells, suggesting that NLRP3-mediated inhibition of STAT1 and NLRC5 is not restricted to tumors (**Extended Data Figs. 7c, d**). Finally, we found NLRP3 upregulation to have no significant impact on IFN-γ receptor tumor surface levels or for caspase-1 activity to influence NLRC5 expression (**Extended Data Figs. 7e, f**).

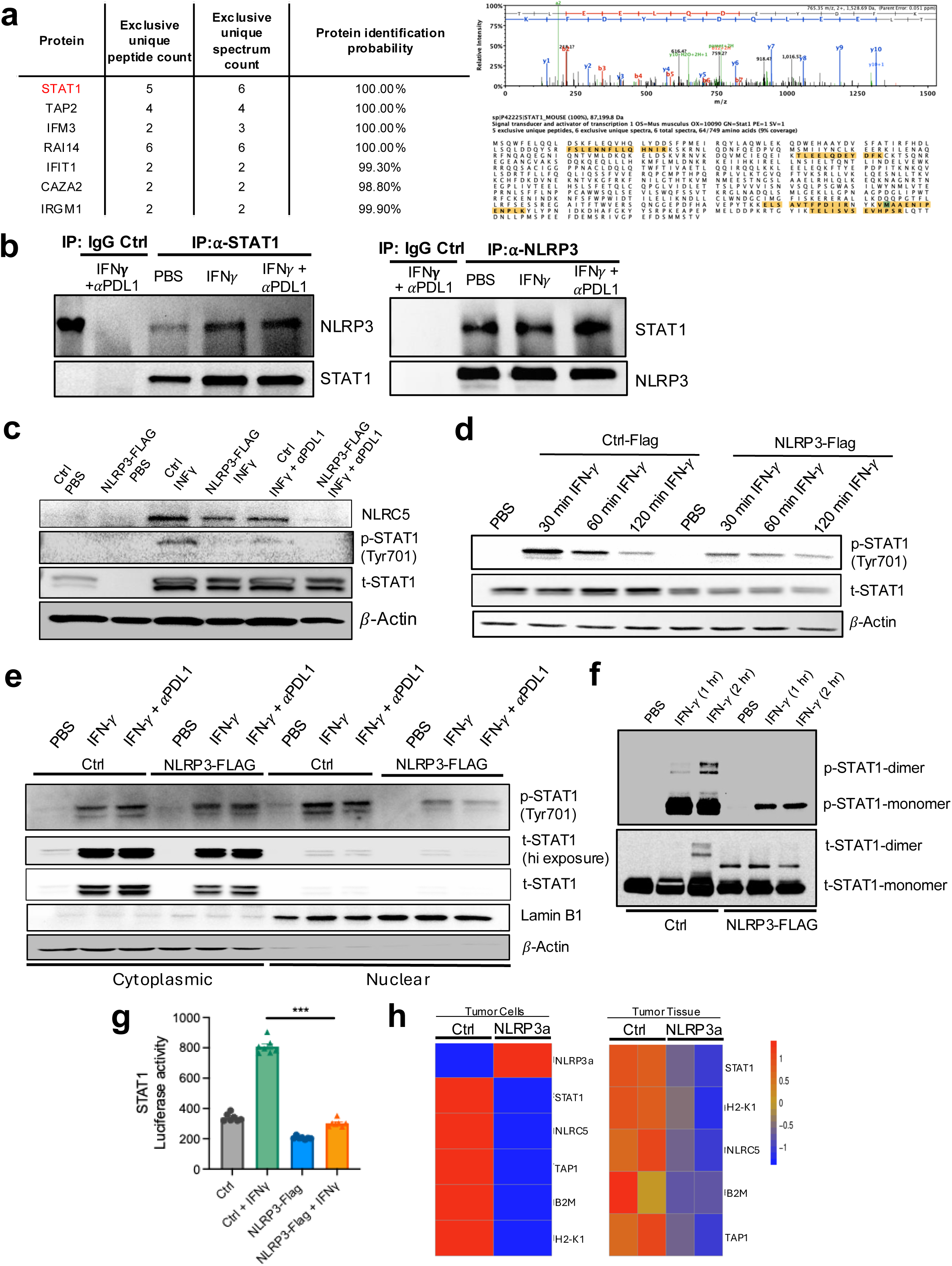

In summary, this work suggests that the NLRP3 inflammasome complexes with STAT1, thereby inhibiting STAT1 dimerization and nuclear translocation. This process suppresses STAT1-dependent transcription of NLRC5 and appears to be the most dominant mechanism inhibiting downstream expression of MHC class I-associated genes in tumor cells.

### NLRP3 Inhibition Reverses STAT1-NLRC5-MHC Class I Suppression and Enhances Response to Anti-PD-1 Immunotherapy in an Orthotopic Model of Gastric Adenocarcinoma

After verifying that NLRP3 activation also suppresses STAT1 signaling in the CIN KPS gastric cancer model, we further confirmed that inhibition of the NLRP3 inflammasome augments nuclear phosphorylated STAT1 levels both *in vitro* and *in vivo* in CIN KPS gastric tumors (**Fig. 7a, Extended Data Figs. 8a-c**). This finding correlated with enhanced MHC class I expression *in vitro*, as well as increased NLRC5 and MHC class I expression by *in situ* tumor tissues following treatment with a NLRP3 inhibitor (**Figs. 7b, c, Extended Data Fig. 8d**). These observations prompted us to explore whether a NLRP3 inhibitor may overcome resistance to anti-PD-1 immunotherapy in a syngeneic subcutaneous CIN KPS gastric cancer model. This study demonstrated NLRP3 inhibitor therapy alone and in combination with anti-PD-1 to suppress tumor progression more effectively than anti-PD-1 monotherapy (**Extended Data Fig. 8e**). Flow cytometry analysis of these resected tumors showed this effect to correlate with diminished PMN-MDSC recruitment and enhanced levels of CD8^+^ T cell infiltration (**Extended Data Fig. 8f**). To evaluate the impact of a NLRP3 inhibitor in a more rigorous model of GE adenocarcinoma, we evaluated NLRP3 inhibitor/anti-PD-1 combination therapy versus anti-PD-1 alone in murine hosts harboring orthotopic CIN KPS gastric cancers. After implanting the CIN KPS gastric cell line by injection into the gastric epithelium, tumors were allowed to progress for 8 days followed by anti-PD-1 immunotherapy every 3 days alone or in combination with the NLRP3 inhibitor, MCC950, dosed every other day (**Fig. 7d**). Consistent with our prior data, NLRP3 inhibition suppressed primary CIN gastric tumor growth while anti-PD-1 immunotherapy alone had minimal effect (**Fig. 7e, Extended Data Fig. 8g**). This study showed anti-PD-1 immunotherapy to enhance tumor-intrinsic NLRP3 signaling activity, an effect that was reversed with the NLRP3 inhibitor, MCC950 (**Fig. 7f**) ^5,31^. Based on both flow cytometry and IHC, we further found NLRP3 inhibition to suppress PMN-MDSC accumulation while enhancing CD8^+^ T cell infiltration into orthotopic CIN gastric tumors (**Fig. 7g, h, Extended Data Fig. 8h**). Cumulatively, these data indicate that the tumor-intrinsic NLRP3 inflammasome contributes to immune evasion via multiple mechanisms and that blocking this pathway may facilitate anti-PD-1 responses in immunotherapy refractory tumors such as GE adenocarcinoma.

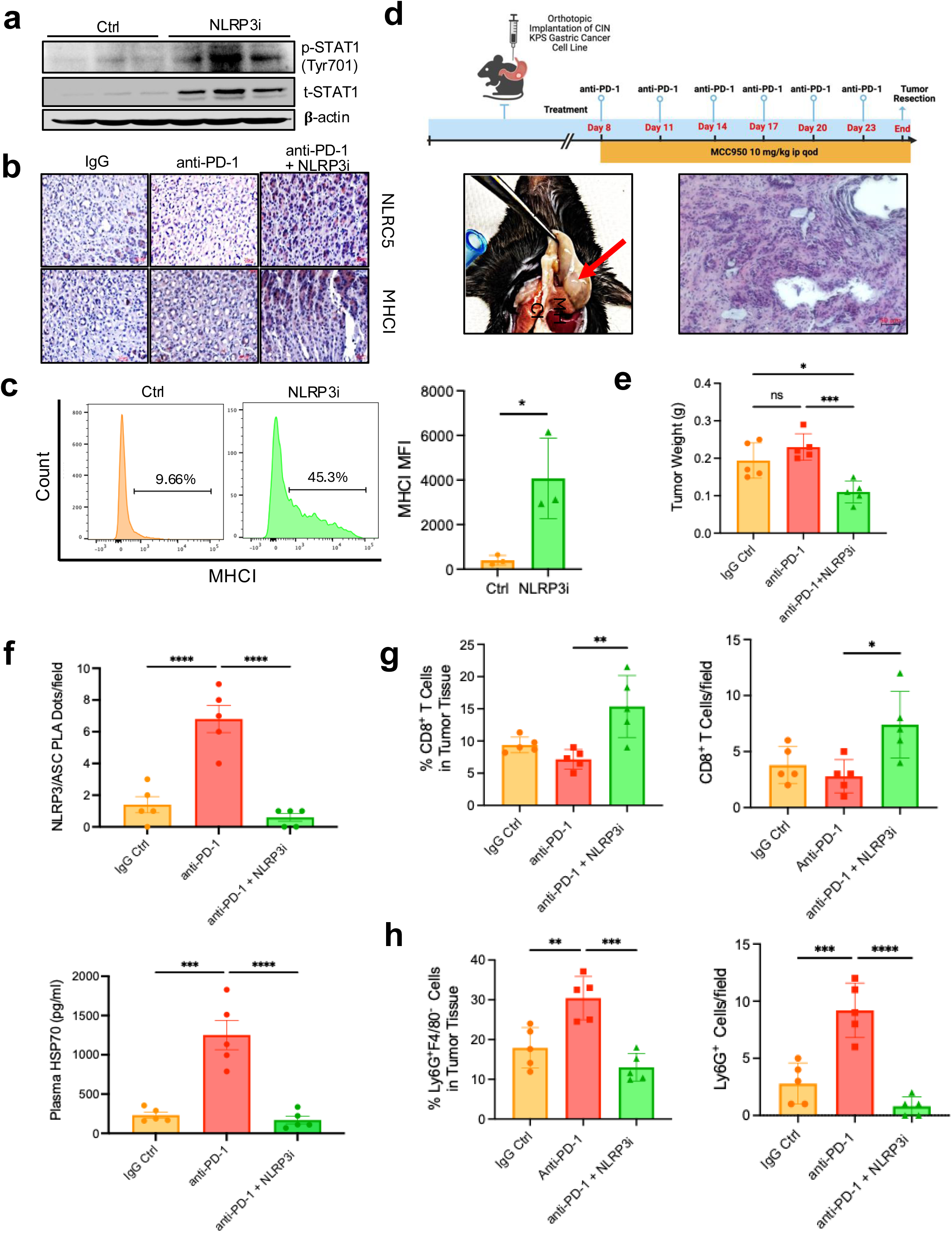

## Discussion

Using an array of pre-clinical tumor models, we have previously reported that the tumor-intrinsic NLRP3 inflammasome triggers the recruitment of PMN-MDSCs into the TME in response to the induction of CD8^+^ T cell activation ^5,6^. We now report that activation of the tumor-intrinsic NLRP3 inflammasome also suppresses NLRC5-mediated transcriptional activation of MHC class I-associated genes, thus impeding CD8^+^ T cell immunosurveillance. Initially observed in GE adenocarcinoma tissue specimens based on immunofluorescence microscopy and spatial transcriptomic studies, this inverse relationship between tumor NLRP3 signaling and NLRC5 expression was also confirmed in human melanoma tissue specimens and then mechanistically verified in studies utilizing human melanoma cell lines as well as in both an orthotopic model of gastric adenocarcinoma and an autochthonous model of melanoma. This work demonstrates that upon oligomerization, NLRP3 binds to STAT1, inhibiting its dimerization, nuclear translocation, and transcriptional activation of NLRC5 ^38^. The current study further suggests that tumor NLRP3 signaling activity, as well as the *NLRP3* genetic amplification level, correlates with resistance to anti-PD-1 immunotherapy based on tumor and plasma specimens derived from two independent cohorts of stage IV melanoma patients (62 patients) as well as smaller cohorts of stage III melanoma patients and advanced GE adenocarcinoma patients (13 and 35 patients, respectively). These findings are further supported by an analysis of three independent transcriptional datasets of advanced melanoma patients treated with anti-PD-1 immunotherapy (98 patients). Given this cumulative data, we propose that this pathway serves as an adaptive resistance mechanism that may contribute to either primary or secondary immunotherapy resistance depending upon the kinetics of its activation.

There has been an extensive body of literature describing the activation mechanisms of the NLRP3 inflammasome in myeloid cell populations and investigators have described two distinct steps involved in this process, including a priming phase which serves to enhance NLRP3 expression levels and an activation phase comprised of a trigger to induce the formation of the multi-molecular NLRP3 inflammasome ^39^. We propose that *NLRP3* copy number gains functionally serve as the priming phase in malignant cells, whereby those tumor cells harboring copy number gains of *NLRP3* are more likely to stimulate the NLRP3 signaling pathway in response to the activation of tumor-infiltrating CD8^+^ T cells. Interestingly, a prior study evaluating somatic mutations and copy number alterations in a series of solid tumors found *NLRP3* copy number gains to be particularly common ^7^. This is further consistent with previous work demonstrating that the 1q44 chromosomal region harboring *NLRP3* to be frequently gained and associated with an inferior prognosis in several solid tumors ^40,41^. While it would seem that *NLRP3* amplification may place tumors at risk for enhanced pyroptotic death, we do not observe this effect and propose that certain tumor-intrinsic alterations are uncoupling pyroptosis from NLRP3 activation. Overall, these findings suggest that further studies are in order to verify the use of *NLRP3* FISH assays to predict immunotherapy outcomes in cancer patients.

Importantly, our prior work has indicated that the activation of this pathway is dependent upon tumor PD-L1 expression ^5^. Indeed, genetic silencing of tumor PD-L1 expression inhibits NLRP3 inflammasome activation and downstream expression of the necessary CXCR2 chemokine gradient required for PMN-MDSC recruitment. The impact of checkpoint inhibitor immunotherapy in oncology has remained limited in many tumor types despite the execution of an array of clinical trials testing combination anti-PD-(L)1 regimens ^42^. One of the primary factors contributing to this barrier in immuno-oncology is the lack of a clinically practical predictive biomarker capable of reliably identifying those tumors that are unlikely to respond to anti-PD-(L)1 regimens ^2^. While PD-L1 IHC-based scoring has been shown to enrich for those tumors more likely to respond to anti-PD-(L)1 immunotherapy, PD-L1 IHC testing often fails to correlate with clinical outcomes ^43^. This has contributed to controversies regarding PD-L1 scoring thresholds for use of anti-PD-(L)1 immunotherapy in certain tumor types, such as in GE adenocarcinoma ^44,45^. Based on our findings, we propose that the tumor-intrinsic NLRP3 inflammasome pathway contributes to checkpoint inhibitor resistance in PD-L1-positive tumors and that tumor-restricted NLRP3 activation markers or the *NLRP3* FISH assay may serve to improve our ability to predict clinical outcomes for candidates of the checkpoint inhibitor immunotherapies.

We developed an unbiased transcriptional signature of tumor-intrinsic NLRP3 inflammasome signaling activity in order to examine the impact of this pathway on clinical response to anti-PD-1 immunotherapy in additional cohorts of melanoma patients. In addition to extending our analysis to larger patient numbers, this approach was taken to allow for a direct comparison with previously published transcriptional signatures developed to predict response to checkpoint inhibitor immunotherapy ^21,22^. Prior studies have developed NLRP3 inflammasome signatures utilized to study the impact of this pathway on tumor biology ^30,46^. However, the approach used to construct these signatures has been problematic in that they often will utilize selected components of the known NLRP3 inflammasome signaling pathway and do not consider feedback inhibition of NLRP3 signaling activity on the expression of genes involved in the pathway. In addition, NLRP3 may have functions that do not involve components of the canonical inflammasome pathway. To overcome these deficiencies, we engineered and transcriptionally profiled an autochthonous *Nlrp3*-silenced BRAF^V600E^-PTEN^-/-^ melanoma model (BRAF^V600E^PTEN^-/-^NLRP3^-/-^) as well as a *Nlrp3*-amplified BRAF^V600E^-PTEN^-/-^ melanoma cell line (BRAF^V600E^PTEN^-/-^NLRP3a). Those genes upregulated in syngeneic BRAF^V600E^-PTEN^-/-^NLRP3a tumors were integrated with genes found to be downregulated in autochthonous BRAF^V600E^-PTEN^-/-^NLRP3^-/-^ tumors. The resulting 48 gene transcriptional signature for tumor-intrinsic NLRP3 signaling activity (NLRP3-tsig) correlates closely with a previously reported transcriptional signature for PMN-MDSCs in both human melanomas and gastric adenocarcinomas, yet is superior to the PMN-MDSC signature in predicting resistance to anti-PD-1 immunotherapy in advanced melanoma patients. Additional studies demonstrate that the NLRP3 activity signature performed favorably versus other published transcriptional signatures in predicting resistance to anti-PD-1 immunotherapy in two additional cohorts of stage IV melanoma patients ^21,22,29^. These results further support the importance of tumor-intrinsic NLRP3 signaling in mediating resistance to anti-PD-1 immunotherapy.

MHC class I downregulation is a well-known mechanism of tumor-mediated immune evasion and the development of anti-PD-1 resistance ^14,15^. Studies have also suggested that inducing tumor MHC class I expression can augment responses to anti-PD-1 immunotherapy ^47^. Once a tumor loses the capacity for MHC class I antigen processing and presentation, there are few pathways that can be leveraged to reverse this effect and there are no therapeutics available to overcome this barrier in the clinic. Herein, we show that systemic pharmacologic inhibition of NLRP3 enhances NLRC5-mediated upregulation of MHC class I expression in tumors *in vivo*, suggesting that NLRP3 inhibition promises to augment other immunotherapy strategies. It is also conceivable that NLRP3-mediated inhibition of MHC class I antigen processing and presentation may contribute to tumorigenesis by interfering with immunosurveillance, serving as a potential mechanistic link between inflammation and carcinogenesis.

Recent studies have demonstrated that chronic IFN-γ stimulation serves to suppress anti-tumor immunity ^48^. We have found that IFN-γ is necessary to stimulate the tumor-intrinsic NLRP3 signaling pathway, raising the possibility that the induction of tumor NLRP3 activation may contribute to the immunosuppressive effects of chronic IFN-γ exposure.

This work suggests that tumor NLRP3 inflammasome activity correlates with enhanced levels of PMN-MDSCs within the TME and resistance to immunotherapy in GE adenocarcinoma patients. These findings were further supported using both genetically engineered syngeneic and orthotopic models of gastric adenocarcinoma, showing that pharmacologic inhibition of NLRP3 significantly suppresses PMN-MDSC recruitment, enhances tumor MHC class I expression, and suppresses tumor progression. Recent clinical studies have demonstrated limited benefit of CXCR2 inhibitors in combination with pembrolizumab in tumor types that typically respond poorly to immunotherapy ^49^. Based on our data, we would conjecture that targeting tumors harboring copy number gains in *NLRP3* are more likely to benefit from this treatment approach. We would further propose that targeting the NLRP3 inflammasome pathway is likely to be more effective than direct CXCR2 inhibition given its additional impact on tumor MHC class I antigen presentation based on the studies presented here.

The treatment arsenal available for patients with GE adenocarcinoma is extremely limited and the prognosis of patients with advanced GE adenocarcinoma remains poor. Given our cumulative data, we propose that clinical trial testing of NLRP3 inhibitors in combination with anti-PD-1 immunotherapy is indicated. Such a study would further provide a platform for verifying our hypotheses that *NLRP3*-amplified GE adenocarcinomas are resistant to anti-PD-1 immunotherapy regimens regardless of their level of PD-L1 expression and that targeting NLRP3 is more likely to be effective as a treatment strategy for *NLRP3*-amplified tumors.

## Methods

### Clinical Samples

Patients provided written informed consent for use of biological specimens on institutional review board (IRB)-approved protocols at Duke Cancer Institute (NCT02694965, NCT03342937) or at Vanderbilt University Medical Center (Institutional Protocol #: 100178). Baseline formalin-fixed paraffin-embedded (FFPE) tissues and plasma specimens were collected from 52 patients with untreated stage IV melanoma prior to initiating anti-PD-1 monotherapy or ipilimumab/nivolumab combination immunotherapy at either Duke Cancer Institute or Vanderbilt University Medical Center (**Supplementary Table 1**). Treatment response at week 12 and every 12 weeks thereafter was determined based on independent radiologic review of computed tomography (CT) imaging using RECIST1.1 (Response Evaluation Criteria in Solid Tumors v1.1) criteria. Patient were considered responders (R) if they demonstrated complete or partial response and considered non-responders (NR) if they demonstrated progressive disease. All patients were monitored for disease recurrence for a total of 5 years. Baseline FFPE tissues and plasma specimens were collected from 13 patients with untreated stage III melanoma prior to initiating adjuvant anti-PD-1 monotherapy at Duke Cancer Institute (**Supplementary Table 2**). Patients underwent CT imaging every 12 weeks and were monitored for disease recurrence for a total of 5 years. Baseline FFPE tissues and plasma specimens were collected from 35 patients with unresectable stage III or stage IV gastroesophageal adenocarcinoma at Duke Cancer Institute prior to initiating pembrolizumab on the IRB-approved KeyLARGO clinical trial (NCT03342937) (**Supplementary Table 4**). Repeat tissue biopsies and plasma specimens were collected on day 8 of treatment prior to initiating capecitabine/oxaliplatin chemotherapy. PFS and best objective response (BOR) were recorded based on day 29 CT imaging followed by subsequent CT imaging every 12 weeks. Assessment of PFS and best overall response rate (BORR) were the primary clinical objectives of this study. All collected tumor tissues were reviewed by a board-certified pathologist prior to analysis.

### Animal Studies

C57BL/6J (C57, H-2^b^) (Stock #: 000664) and B6.CgBraf^tm1Mmcm^Pten^tm1Hwu^Tg(Tyr-cre/ERT2)13Bos/BosJ (Braf^V600E^Pten^−/−^, H-2^b^) (Stock #: 012328) mice were purchased from Jackson Labs. C57BL/6-Nlrp3 ^tm1.1Mrl^ (NLRP3^flox/flox^, Stock #: 12809) were obtained from Taconic. The NLRP3^flox/flox^ mice were crossed with B6.Cg-*Braf^tm1Mmcm^ Pten^tm1Hwu^* Tg(Tyr-cre/ERT2 H-2^b^)13Bos/BosJ transgenic mice to produce a melanoma tissue-specific *Nlrp3* conditional knockout. NLRP3 silencing was verified by PCR and immunoblotting. Mice were given 100 µl tamoxifen (Sigma-Aldrich, CAS# 10540-29-1, 20 mg/mL) through intraperitoneal (i.p.) injection daily for 5 days, with a total dose of 75 mg/kg [47]. All experimental groups consisted of randomly chosen littermates of both sexes, aged 6 to 10 weeks, from the same strain. All animal procedures followed protocols approved by the Institutional Animal Care and Use Committee at either Duke University Medical Center or the University of North Carolina at Chapel Hill.

### Autochthonous Tumor Studies

B6.Cg-*Braf^tm1Mmcm^ Pten^tm1Hwu^* Tg(Tyr-cre/ERT2 H-2^b^)13Bos/BosJ transgenic mice were subdermally injected with 4-Hydroxytomoxifen (4-HT) (Sigma-Aldrich, H6278-50MG CCF, 38.75 µg/mouse) to induce primary melanoma development at the base of the tail. Mice were randomly assigned to treatment cohorts until tumor volumes reached 64 mm^3^ ^50–52^. For select experiments, mice were treated with the following agents: NLRP3 inhibitor, MCC950 (Invivogen, inh-mcc) 10 mg/kg i.p. injection every other day (qod), anti-PD-1 antibody (BioXCell, BE0146) or rat IgG2a isotype control (BioXCell, BE0089) at 200 µg i.p. every 3 days, anti-Ly6G antibody (BioXCell, BE0075-1), initial dose at 200 μg/mouse followed by 100 μg/mouse/day x 2. Primary tumor volumes were monitored by orthogonal caliper measurements every 3 days. Tumor volume was calculated according to the formula: cm^3^ = [(length, cm) x (width, cm)^2^]/2.

### Syngeneic and Orthotopic Tumor Studies

BRAF^V600E^PTEN^−/−^, BRAF^V600E^PTEN^−/−^NLRP3a, and BRAF^V600E^PTEN^−/−^NTC control cell lines (0.5 x 10^5^ to 1 x 10^6^ cells) were implanted by subcutaneous injection into the base of the tail or chest of syngeneic C57BL/6 mice (NTC, non-target control). Chromosomal instability (CIN) Kras^G12D/+^Tp53^R172H/+^Smad4^-/-^ (KPS) gastric cancer cells (0.5 × 10^6^ cells) in 50 µl of cell suspension with 50% Matrigel were injected either orthotopically into the subserosa of the gastric wall or implanted subcutaneously in syngeneic C57BL/6 mice (0.5 x 10^5^ to 1 x 10^6^ cells) ^36^. Subcutaneous tumor growth was monitored using caliper measurements every 3 days and treatment was initiated when tumor volumes reached either 64 mm³ or 90 mm³, depending on the study. Surgical procedures were performed under a small animal heat lamp and post-surgery recovery was monitored closely. Surgical wounds were managed with surgical clips and topical iodine solution. Treatment consisting of either anti-PD-1 (200 µg i.p. every 3 days), MCC950 (10 mg/kg i.p. injection qod), or both was initiated on day 8 post-implantation. Orthotopic tumor growth was measured based on caliper measurements and stomach weight on day 25.

### Cell Lines and Culture Conditions

BRAF^V600E^PTEN^−/−^NLRP3a and BRAF^V600E^PTEN^−/−^NTC cell lines were engineered using the CRISPR amplification technique as previously described ^31^. The NLRP3 and control CRISPRa plasmids (Santa Cruz, sc-432122-ACT) were packaged into a lentiviral vector in HEK293T cells as previously described [53]. Clones were selected using a combination of antibiotics, including puromycin (Santa Cruz, sc-108071), hygromycin B (Santa Cruz, sc-29067), and blasticidin S HCL (Santa Cruz, sc-495389). The BRAF^V600E^PTEN^−/−^ (male, BPD6) melanoma cell line was derived from the transgenic BRAF^V600E^PTEN^−/−^ melanoma model as previously described ^50,53^. The BRAF^V600E^PTEN^−/−^NLRP3^KD^ and BRAF^V600E^PTEN^−/−^NTC cell lines were derived from the BRAF^V600E^PTEN^−/−^ cell line as previously described [54]. Stable cell lines were selected based on puromycin resistance (Sigma-Aldrich, P8833). The BRAF^V600E^PTEN^−/−^NLRP3-F and the BRAF^V600E^PTEN^−/-^Flag control cell lines were generated using the pcDNA3-N-Flag-NLRP3 (Addgene, 75127) and pcDNA3-Flag (Addgene, 20011) plasmids, respectively. The BRAF^V600E^PTEN^−/−^STAT1-GFP cell line was created using the pLenti-C-STAT1-mGFP-P2A-Puro vector (Addgene, 199345). Drs. Therese Seidlitz and Daniel E Stange (Carl Gustav Carus Unversitätsklinikum, Dresden, Germany) generously provided the CIN KPS gastric cell line (Kras^G12D/+^ Tp53^R172H/+^Smad4^-/-^). All BRAF^V600E^PTEN^−/−^ cell lines were maintained at 37°C in Dulbecco’s Modified Eagle Medium (DMEM, Invitrogen) with 2 mM L-glutamine, supplemented with 10% fetal bovine serum (FBS) and 100 units/ml penicillin. The CIN KPS cell line was cultured in advanced DMEM medium with 10% FBS, glutamine, and 1x B57 supplement. For select *in vitro* experiments, cell lines were treated with IFN-γ (100 ng/ml, BioAbChem, 42-IFNg), anti-PD-L1 antibody, clone 10F.9G2 (1 to 2 µg/ml, BioXCell, BE0101), or MCC950 NLRP3 inhibitor (2.5 µM to 10 µM, Invivogen, inh-MCC). All cell lines were screened annually for mycoplasma contamination by the Duke University and University of North Carolina Tissue Culture Facilities.

### RNA Isolation and qrt-PCR Analysis

Tissue specimens were harvested from tumors and stored in RNAlater at –80°C. Tissues were lysed in RLT buffer by using a gentleMACS dissociator (Miltenyi Biotec) and cell lines were lysed in RLT buffer and stored at –80°C. Total RNA was isolated with the RNeasy Plus Mini Kit (Qiagen, 74134) and quantified using a NanoDrop. RNA (500 ng to 1000 ng) was used for cDNA synthesis with the iScript Reverse Transcription Supermix (BioRad, 1708841). qRT-PCR was performed on an ABI7500 Real-Time PCR system (Life Technologies). All qrt-PCR reactions were performed using validated primers (**Supplementary Table 6**) and SsoAdvanced Universal SYBR Green Super Mix (BioRad, 1725271) or SsoAdvanced Universal Probes Supermix (BioRad, 1725281). All data were normalized to β-actin expression and relative gene expression was quantified using the 2^-ΔΔCt^ method.

### Western Blot Analysis

Tissues or cells were lysed in NP40 lysis buffer (Sigma-Aldrich) supplemented with a complete protease inhibitor and phosphatase inhibitor (Roche). After lysing in Laemmli sample buffer, equal volumes of lysates were separated using 10% or 15% SDS–PAGE and then transferred to a PVDF membrane (Bio-Rad Laboratories Inc., 162017). After blocking for 30 minutes in tris-buffered saline (TBS) containing 0.1% Tween-20 and 5% milk, the membranes were probed with various primary antibodies followed by horseradish peroxidase (HRP)-conjugated secondary antibodies. Primary antibodies included: Anti-β-actin mouse monoclonal (clone c4, 1:3000, Santa Cruz Biotechnology, sc-47778), anti-NLRP3 rabbit monoclonal (clone D4D8T, 1:1000, Cell Signaling, 15101S; clone EPR23073-96, 1:1000, Abcam, 270499), anti-Caspase-1-p20 mouse monoclonal (clone Casper-1, 1:500, Adipogen, AG-20B-0042-C100), anti-HSP70 mouse monoclonal (clone C92F3A-5, 1:1000, Santa Cruz Biotechnology, sc-66048. anti-NLRC5 mouse polyclonal (clone IN113, 1:500, Adipogen, AG-25B-0038-C100). Anti-STAT1 rabbit monoclonal (clone D1K9Y, 1:1000, Cell Signaling, 14994S), anti-P-STAT1 rabbit monoclonal (clone 58D6, 1:1000, Cell Signaling, 9167S), and anti-Lamin B1 mouse monoclonal (clone B-10, Santa Cruz Biotechnology, SC-374015). Immunoreactivity was visualized using a chemiluminescence substrate (Thermo Fisher Scientific, 34095/34075) and imaged with a ChemiDoc XRS+ System (Bio-Rad).

### Co-immunoprecipitation Studies

Cells were lysed using IP lysis buffer (Thermo Fisher, 87787). Either Anti-NLRP3 antibody (Abcam, 270449) or Anti-STAT1 (Cell Signaling, 14994S), along with IgG isotype (Cell Signaling, 3900), was conjugated with Dynabeads Protein A beads (Thermo Fisher, 10006D). These conjugates were added to the lysate and processed according to the manufacturer’s protocol. The NLRP3-bound and isotype-bound beads were pulled down using magnetic isolation techniques. After washing, proteins from the beads were eluted and analyzed by Western blot, then probed for either Anti-STAT1 (Cell Signaling, 14994S) or Anti-NLRP3 antibody (Abcam, 270449).

### Liquid Chromatography–Tandem Mass Spectrometry

Target proteins were immunoprecipitated using magnetic beads coated with anti-NLRP3 or isotype control antibodies. Beads were washed 5 times with 50 mM ammonium bicarbonate, followed by digestion with 20 µL of 20 ng/µL trypsin overnight. An additional 20 µL of trypsin was added, and supernatants were recovered after 4 hours. Digests were adjusted to 70 µL with 0.1% TFA and then subjected to Stagetip cleanup. After lyophilization, peptides were resuspended in 30 µL, and 5 µL of each sample was analyzed using a 90-minute FAIMS-LC-MS/MS method on Thermo Fusion Lumos MS at the Duke Proteomics and Metabolomics Shared Resource. Data were processed using FragPipe 16 and visualized in Scaffold v5.3.4 (Portland, OR).

### ELISA

Human plasma HSP70 levels were measured in a blinded fashion using the human HSP70 DuoSet assay (R&D Systems, DY1663). All ELISAs were performed according to the manufacturer’s protocol.

### Flow Cytometry Analysis

Cells (1 to 2 x 10^6^) were diluted in 150 μl flow buffer per well of V bottom culture plates, incubated with Live/Dead Fixable Violet Dead Cell Stain Kit (0.5 μl/1 x 10^6^ cells, Thermo Fisher Scientific, L34964A) on ice for 20 minutes, washed twice, and incubated with Fc Block (anti-CD16/CD32, 2 µg/mL) before staining with appropriate conjugated antibodies. Cells were washed, resuspended in 2% paraformaldehyde for 15 minutes at 4°C, and analyzed using a BD FACSCanto flow cytometry system. Compensation was performed using fluorescence minus one (FMO) controls. Cells were characterized using the following combinations of cell surface markers after gating on viable single cell populations. All antibodies were obtained from commercial vendors and utilized at 1 μg / 1 x 10^6^ cells: Anti-Mouse CD11b, phycoerythrin (PE)-conjugated, clone: MIH5 (BD Pharmingen, 558091), Anti-Mouse Ly6G-GR1, fluorescein isothiocyanate (FITC)-conjugated, clone: RB6-8C5 (BD Pharmingen, 5532127), Anti-Mouse Ly6G, FITC-conjugated, clone: 1A8 (BioLegend,127605), Anti-Mouse Ly-6G, FITC-conjugated, clone: 1A8-Ly6g (Thermo Fisher, 11-9668-82), Anti-Mouse F4/80, APC-conjugated, clone: BM8 (BD Pharmingen, 560408), Anti-Mouse CD45, PerCP-Cy5.5-conjugated, clone: 145-2C11 (BD Pharmingen, 551163), Anti-Mouse CD90.2, FITC-conjugated, clone: 53-2.1 (BD Bioscience, 553003), Anti-Mouse CD31, clone: 390 (Bio legend 102406), FITC-conjugated, clone: 53-2.1 (BD Bioscience, 553003) Anti-Mouse CD8α, brilliant violet (BV) 510-conjugated, clone: 53-6.7 (BD Pharmingen, 563068), Anti-Mouse CD3ε, PerCP-Cy5.5-conjugated, clone: 145-2C11 (BD Pharmingen, 551163), Anti-Mouse CD8α, APC-conjugated, clone: 53-6.7 (BD Bioscience, 553035), Anti-Mouse Ly6C, PE-Cy7-conjugated, clone: AL-21 (BD Pharmingen, 560593), Anti-Mouse CD119, PE-conjugated, clone: 2E2 (Thermo Fisher,12-1191-82). Cell populations were characterized based on the following marker profiles: Ly6G+ PMN-MDSCS: CD45^+^CD11b^+^Ly6G^hi^Ly6C^lo^F4/80^-^; Tumor cells : CD45^-^CD90.2^-^CD31^-^; tumor-associated macrophages: CD45^+^CD11b^+^Ly6G^-^F4/80^+^. FACS was performed using a Beckman Astrios Cell Sorter at cell densities of 1 x 10^6^ cells/mL in 1% RPMI supplemented with penicillin/streptomycin. Flow cytometry data was analyzed using FlowJo software v10.3.

### Immunohistochemistry Analysis

Paraffin sections (5-μm) from human GE adenocarcinoma and mouse gastric cancer tissues were processed using standard protocols for immunohistochemistry (IHC) and immunofluorescence (IF) staining. Tissues were permeabilized by incubation in 0.4% Triton-X in TBS for 20 minutes. The following primary antibodies were incubated for 12-18 hours at 44°C: anti-NLRC5, rabbit polyclonal (clone: 84166, 1:200, Novus Biologicals, NBP3-17901), anti-NLRC5, rabbit polyclonal (clone: AB_2900294, 1:100, ThermoFisher, PA5-115659), anti-pan-MHC class i, rabbit monoclonal (clone: E8E7N, 1:100, Cell Signaling, 76828). Pan-MHC class I and NLRC5 expression levels were analyzed using ImageJ software based on six to eight random fields averaged per section across three to four sections per specimen.

### Spatial Transcriptomics

Human GE adenocarcinoma FFPE tissue specimens were analyzed on the Nanostring GeoMx digital spatial profiling (DSP) platform (Bruker, Seattle, WA) at the UNC Pathology Services Core (PSC) Facility. FFPE tissue samples were prepared on positively charged slides. Pre-treatment was carried out using the Bond III Automated IHC Staining System (Leica Biosystems, Deer Park, IL) and the following primary antibodies: anti-CD8, rabbit monoclonal (Cell Marque, 108R-16) 1:500 for 1 hr; anti-pan-CK, mouse monoclonal (Leica Biosystems, AE1/AE3-601-L-U) 1:300 for 30 mins; anti-ELA2, mouse monoclonal (NovusBio, MAB9167AF647) 1:600 for 1 hr. Tyramide signal amplification was performed using Cy3, Alexa488, and Cy5 fluorophores, respectively. Tissues were dewaxed in Bond Dewax solution (Leica Biosystems, AR9222) and hydrated in Bond Wash solution (Leica Biosystems, AR9590). Post-fixation was accomplished using 10% Neutral Buffered Formalin (Electron Microscopy Science, 15740-04). Heat-induced antigen retrieval was performed in Bond-Epitope Retrieval solution 2 pH 9.0 (Leica Biosystems, AR9640) for 20 min, enzymatic retrieval with Bond Enzyme Pre-treatment Kit (Leica Biosystems, AR9551) at 0.1ug/mL for 15 min. Human Whole Transcriptome Atlas probe was hybridized overnight, slides were washed twice in fresh 2X saline-sodium citrate (SSC, Sigma-Aldrich, S6639-1L) and loaded onto the GeoMx DSP. A pan-cytokeratin IF marker was used to identify epithelial tumor tissue while CD8 and elastase IF markers were used to assess tumor-infiltrating CD8^+^ T cell and PMN/neutrophil cell populations, respectively. Six regions of interests (ROI) were selected at 20X magnification for each tissue slide based on the abundance of each of these markers. UV light was used to cleave oligos that were then collected in a 96-well plate for library preparation and were sequenced on an Illumina NextSeq2000 by the UNC High-Throughput Sequencing Facility (HTSF). All transcriptional data was analyzed using GeoMx DSP Data Analysis software. Following data normalization, differential expression analysis was performed using the ShinyGo gene-set enrichment tool ^33^.

For quality control, barcode yields were evaluated to ensure adequate RNA capture with negative control probes defining background signal. Probes were removed globally from the dataset if either of the following was true: 1) the geometric mean of that probe’s counts from all segments divided by the geometric mean of all probe counts representing the target from all segments is less than 0.1, or 2) the probe is an outlier according to the Grubb’s test in at least 20% of the segments. Read depth per ROI, alignment rates, unique molecular identifier (UMI) counts, and gene detection above background were reviewed. Outlier ROIs were flagged and removed prior to downstream analysis. The Limit of Quantification (LOQ), defining the minimum expression level at which a transcript can be reliably and confidently distinguished from background noise in each ROI was determined and segments with exceptionally low signal were filtered out.

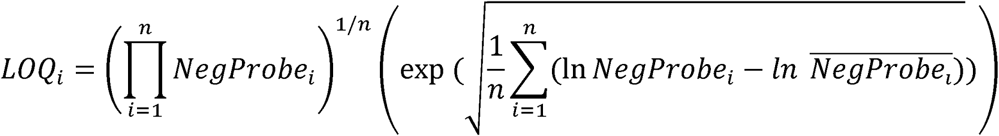

where n is the total number of probes. After quality control review, 81 samples (ROIs) x 1304 genes passed. Finally, dimensionality reduction (PCA) and normalization techniques (Q3 normalization) were used to identify potential outliers, and none were observed.

### Proximity Ligation Assay

Proximity ligation assays (PLAs) were performed as previously described ^31^. In brief, human tumor tissues were deparaffinized, rehydrated, subjected to antigen retrieval, permeabilized in 0.4% Triton-X in TBS for 20 min, and blocked using 0.1% BSA in TBS and 0.05% Tween solution for 30 minutes at room temperature. Slides were incubated overnight with human NLRP3 (aa 540-689) antibody (1:200, 0.2 mg/mL, R&D Systems, AF7010) and anti-human ASC mouse monoclonal antibody (1:200, 0.2 mg/mL, Santa Cruz Biotechnology, sc-514414) at 4°C. A mixture of 1x DuoLink in situ PLA probe anti-mouse PLUS and 1× DuoLink PLA probe anti-goat MINUS (Sigma-Aldrich, DU092001-30RXN) was added to the section and incubated for 1 hour at 37°C. Ligase solution was added to each sample for an additional 30 minutes followed by an amplification solution (35 μl) and incubated for 100 minutes. Finally, mounting medium with 4’,6-diamidino-2-phenylindole (DAPI) was added and a cover slip was placed. Images were captured using a SP5 Leica confocal microscope. ImageJ software was used to quantify fluorescent dots in a blinded fashion based on three fields per tissue section at 40X (averaged over two tissue sections per tissue sample).

### Fluorescence *In Situ* Hybridization

FFPE tissue slides were deparaffinized and pre-treated with citric acid buffer, then digested with pepsin. After washing, the tissue slides were denatured and hybridized with a *NLRP3* FISH probe (Empire Genomics, NLRP3-10-GR) at 75°C for 10 minutes and 37°C for 16 hours and then counterstained with DAPI. A centromere probe for chromosome 1 (CEP1) served as an internal control (Empire Genomics, 1Q-10-O-A). The slides were examined under an SP8 Leica Confocal Microscope and the NLRP3:CEP1 amplification ratio was quantified using the LASX suite in a blinded fashion. NLRP3 and CEP1 ratios were quantified using established guidelines for the enumeration of HER2 amplification ^54^. The total number of NLRP3 counts were divided by the total number of CEP1 counts in at least 20 non-overlapping nuclei.

### RNAseq and Tumor-intrinsic NLRP3 Transcriptional Signature Analysis

Flash-frozen tumors were placed in M-tubes (Miltenyi, 130-093-236) with RLT PLUS (QIAGEN, 1053393) containing β-mercaptoethanol (VWR, VWRV0482-250ML) and homogenized using the gentleMACS automated homogenization system. RNA was extracted from the tumors with the RNeasy Micro Kit (Qiagen, 74004). Messenger RNA was isolated from total RNA using poly-T oligo-attached magnetic beads. Subsequently, cDNA was synthesized. The library was prepared with the NEBNext Ultra II RNA Library Prep Kit. Quantified libraries were pooled and sequenced on the Illumina NovaSeq 6000 system based on their effective concentration and data requirements. Each sample yielded twenty million reads at PE150 depth.

Based on the bulk RNA-seq data from the whole tumor, common elements of the gene set were identified between the most significantly upregulated genes in *Nlrp3*-amplified tumors and the most significantly downregulated genes in *Nlrp3*-silenced tumors. This gene set was cross-referenced with detectable transcripts in human melanoma tissues based on the TCGA in order to generate a 48-transcript NLRP3-tsig signature. Publicly available RNA-seq datasets were filtered for samples with baseline tissue specimens and restricted to only naive stage IV melanoma patients receiving anti-PD-1 monotherapy. The raw RNA-seq data from the patient’s samples was initially normalized by converting it to transcripts per million. Next, the ssGSEA score was computed for each sample using the generated NLRP3-tsig signature. Finally, a ROC curve was created based on these ssGSEA scores and the binary treatment response outcomes (R or NR). Patient were considered responders (R) if they demonstrated complete response, partial response, or stable disease, while they were considered non-responders (NR) if they demonstrated progressive disease.

### TCGA Data Analysis

The TCGA data were accessed and visualized through cBioPortal and GEPIA2 ^20,55^.

### Statistics

Unpaired t-tests were used to compare mean differences between control and treatment groups. One- or two-way ANOVAs followed by Tukey’s multiple comparisons test or Sidak’s multiple comparisons test, respectively, were performed to analyze data containing three or more groups. Various parameters were correlated based on Pearson correlation coefficient analysis. PFS and OS in patients with stage IV melanoma undergoing anti-PD-1 immunotherapy was analyzed using a log-rank test. All tests were two-sided with *P* values reported. A *P* value of less than 0.05 was considered significant. All quantitative data are presented as the mean ± SEM. Specific statistical tests are reported in the Figure Legends. GraphPad Prism 10 was used for all statistical analyses.

## Data Availability

All data produced in the present study are available upon reasonable request to the authors following publication.

## Acknowledgements.

We would like to thank Drs. Therese Seidlitz and Daniel E Stange (Carl Gustav Carus Unversitätsklinikum, Dresden, Germany) for generously providing the Anxa10-CreER^T2^;Kras^G12D/+^;Tp53^R172H/+^;Smad4^fl/f^ CIN gastric adenocarcinoma cell line for these studies. We thank Dr. William Jeck (Duke University, Durham, NC) for his generous review of the GE adenocarcinoma tissue biopsy specimens. We would like to thank Edison Floyd and Gabriela De La Cruz in the UNC Pathology Services Core (PSC) Facility at UNC Lineberger Comprehensive Cancer Center for their expert assistance with the Nanostring GeoMx spatial transcriptomic studies. The UNC PSC is supported in part by an NCI Center Core Support Grant (P30CA016086). Finally, we are grateful for the assistance of Hongwei Liu and Dr. Corbin Jones in the UNC Bioinformatics and Analytics Research Collaborative (BARC).

## Author Contributions

B.T. and B.A.H. conceptualized and managed the project. B.T. or N.Y. performed or directed all experiments. K.V., Y.N., L.C., E.P.C. supported Western blot, qrt-PCR, and mouse studies. N.C.D. and K.M. supported the bioinformatic studies. B.A.H., D.J., J.B., E.B., H.U., and J.C.S. provided clinical resources for the project. B.A.H. wrote the manuscript. B.A.H., B.T., N.C.D., and M.P.P. reviewed and edited the manuscript.

