## Supplementary Figures_BH for "Tumor *NLRP3* Amplification Promotes Immunotherapy Resistance by Suppressing MHC Class I Expression"

### Extended Data Fig. 1.

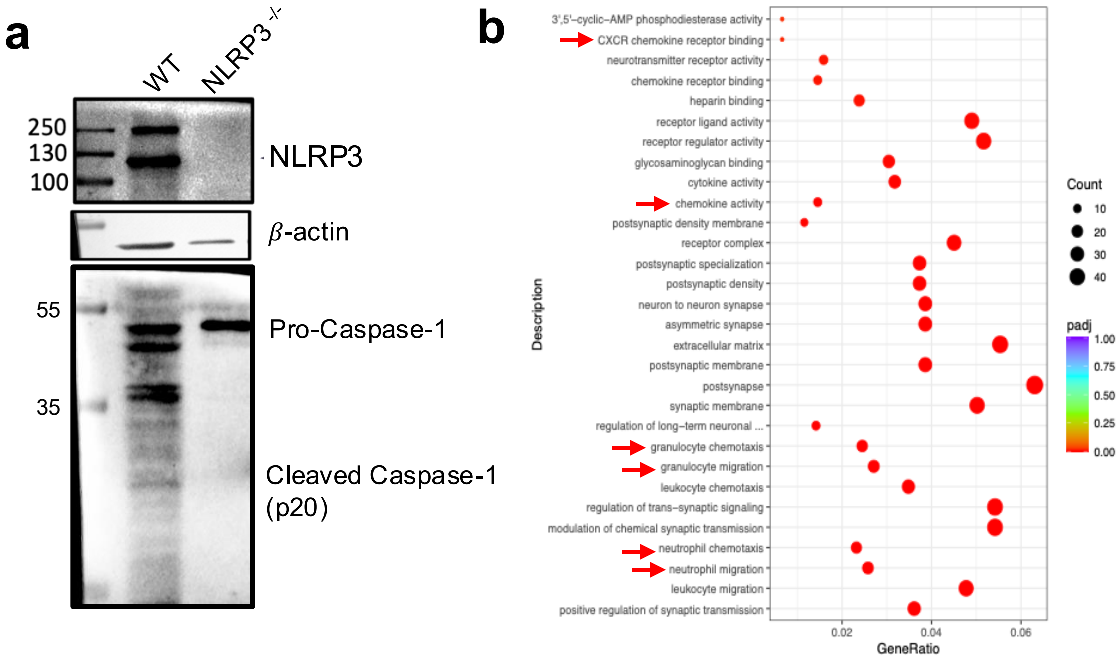

### Extended Data Fig. 2.

a

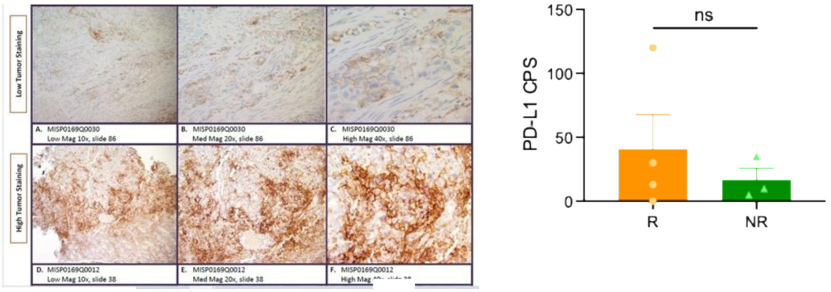


b

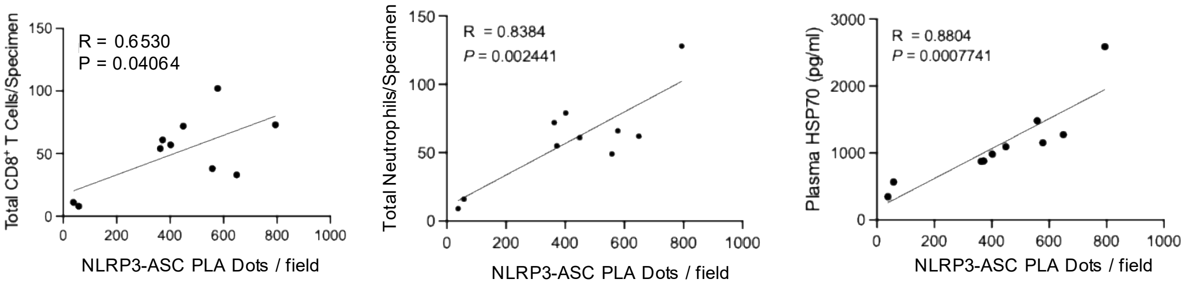

### Extended Data Fig. 3.

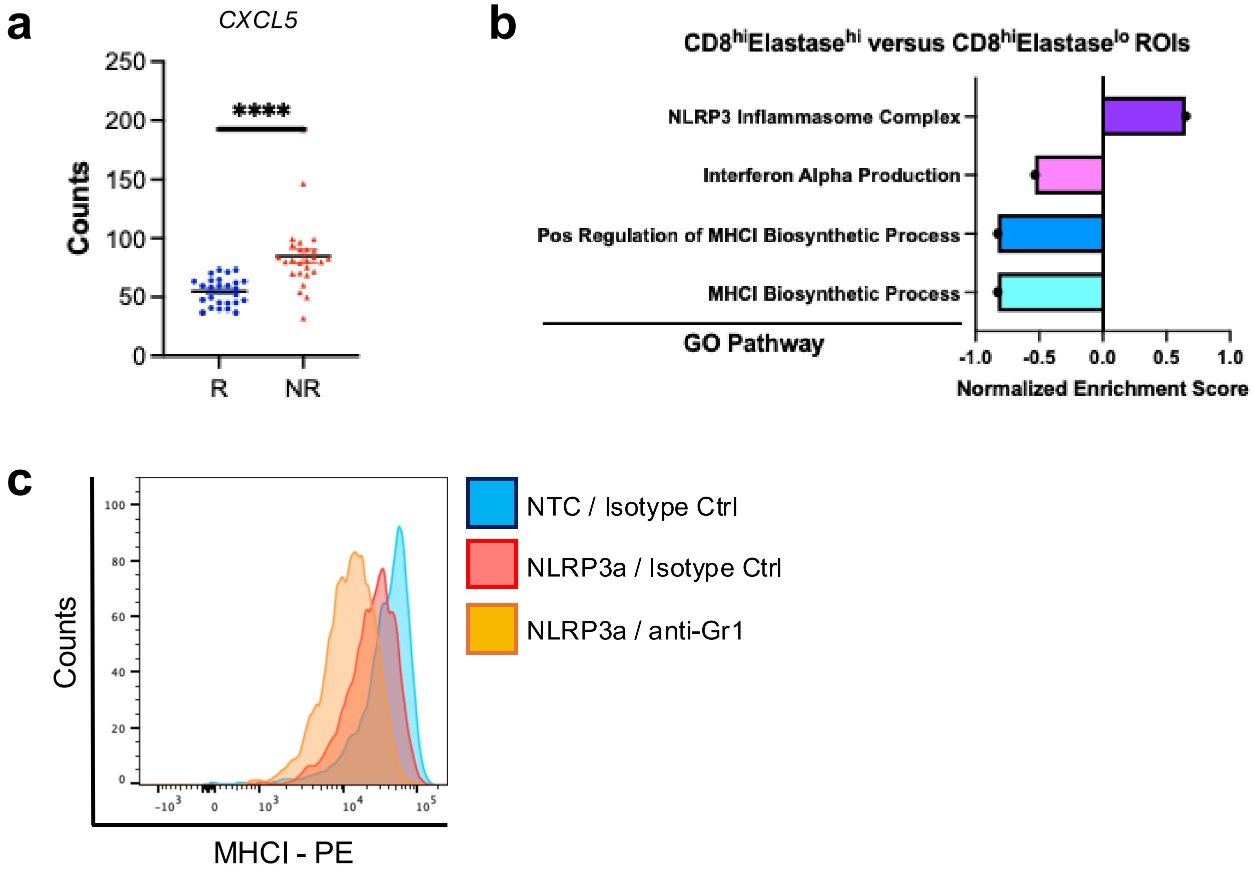

### Extended Data Fig. 4.

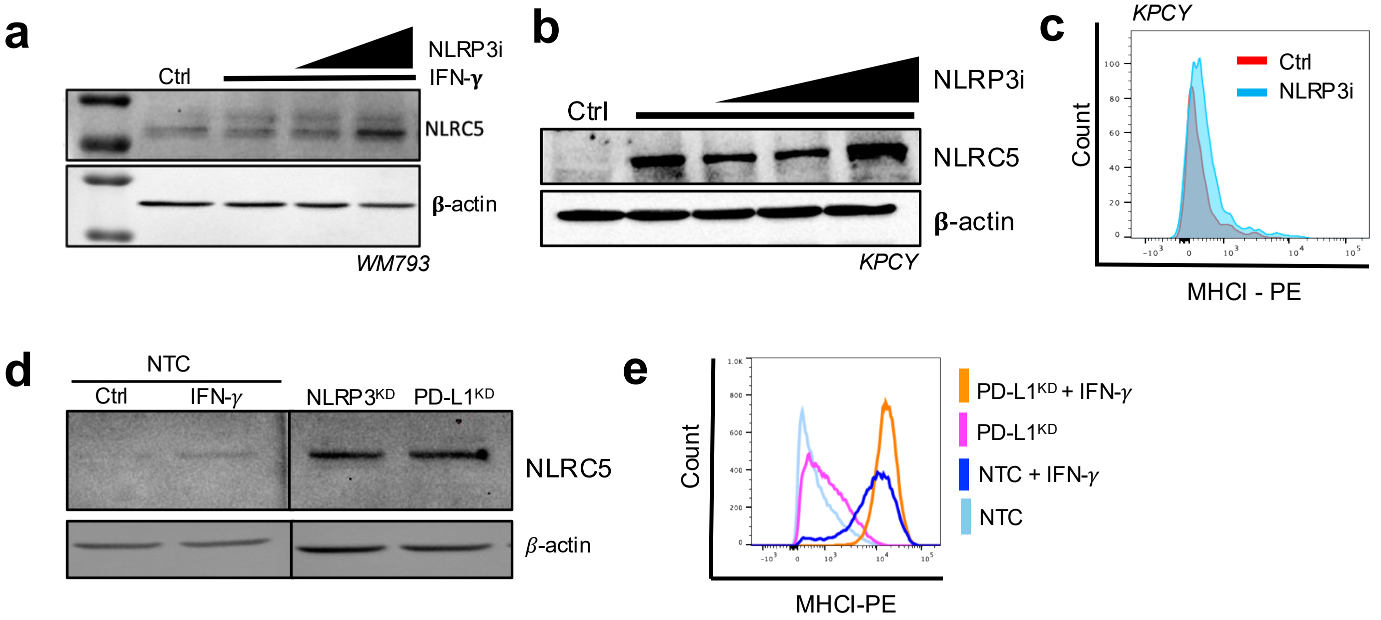

### Extended Data Fig. 5.

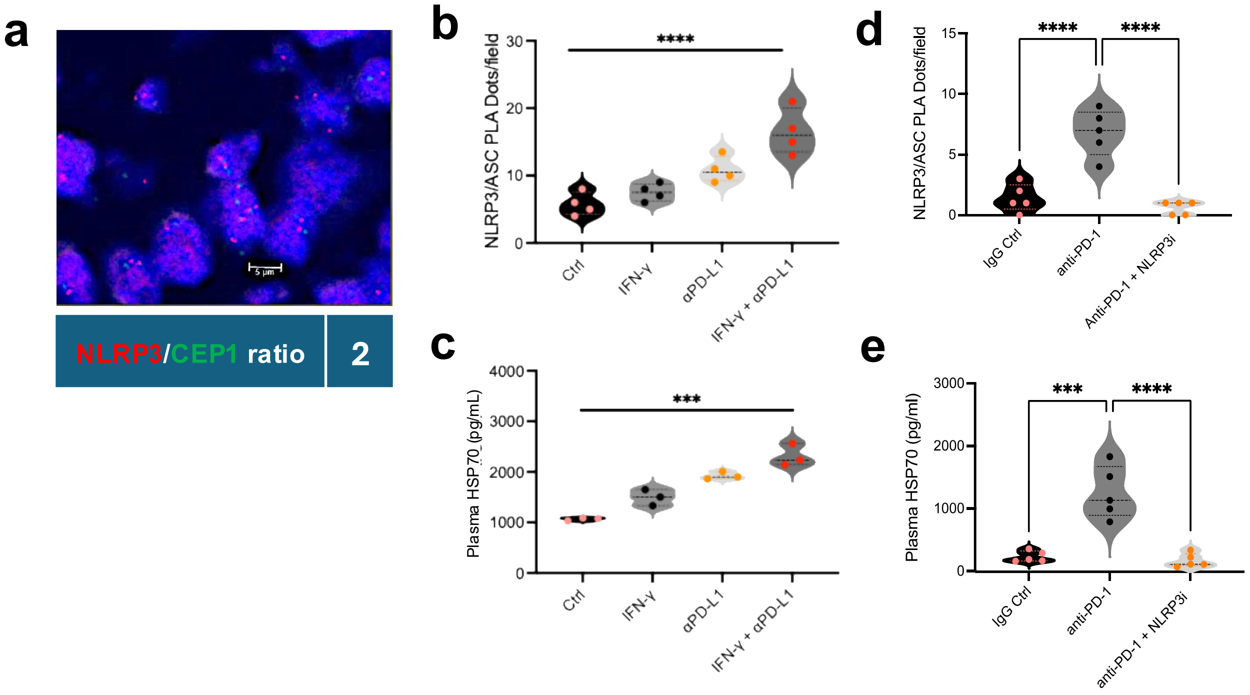

#### Extended Data Fig. 6.

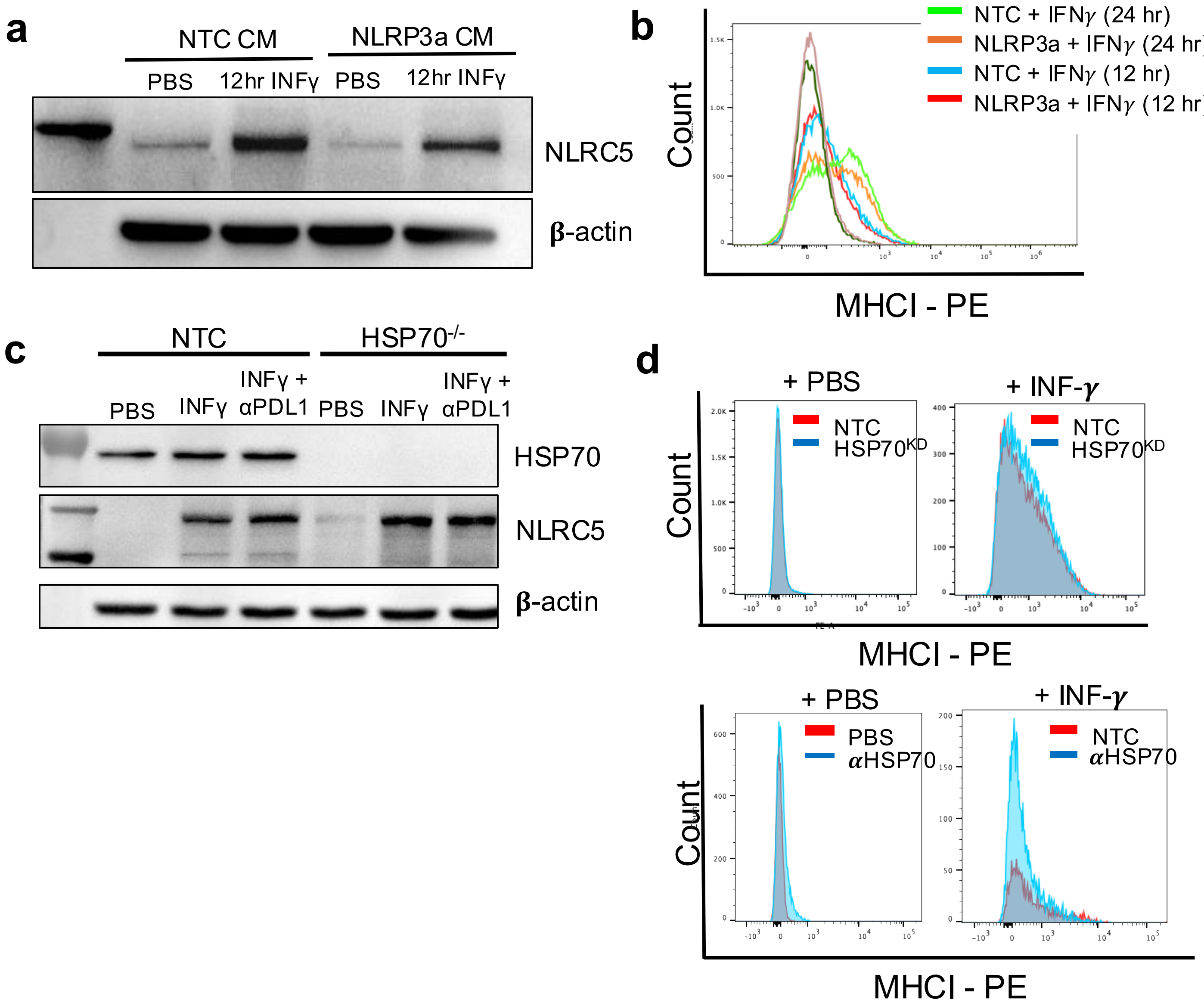

### Extended Data Fig. 7.

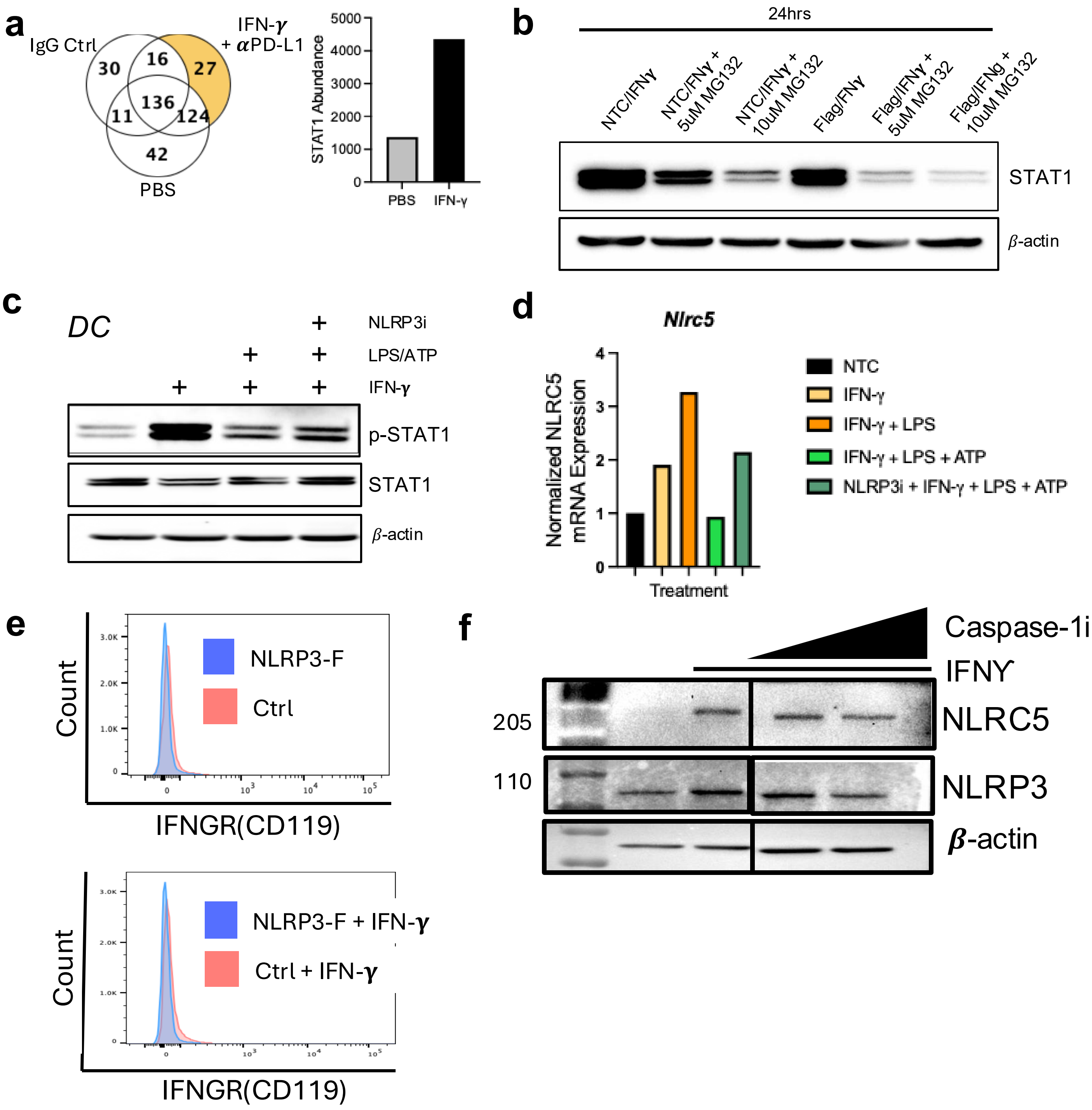

### Extended Data Fig. 8.

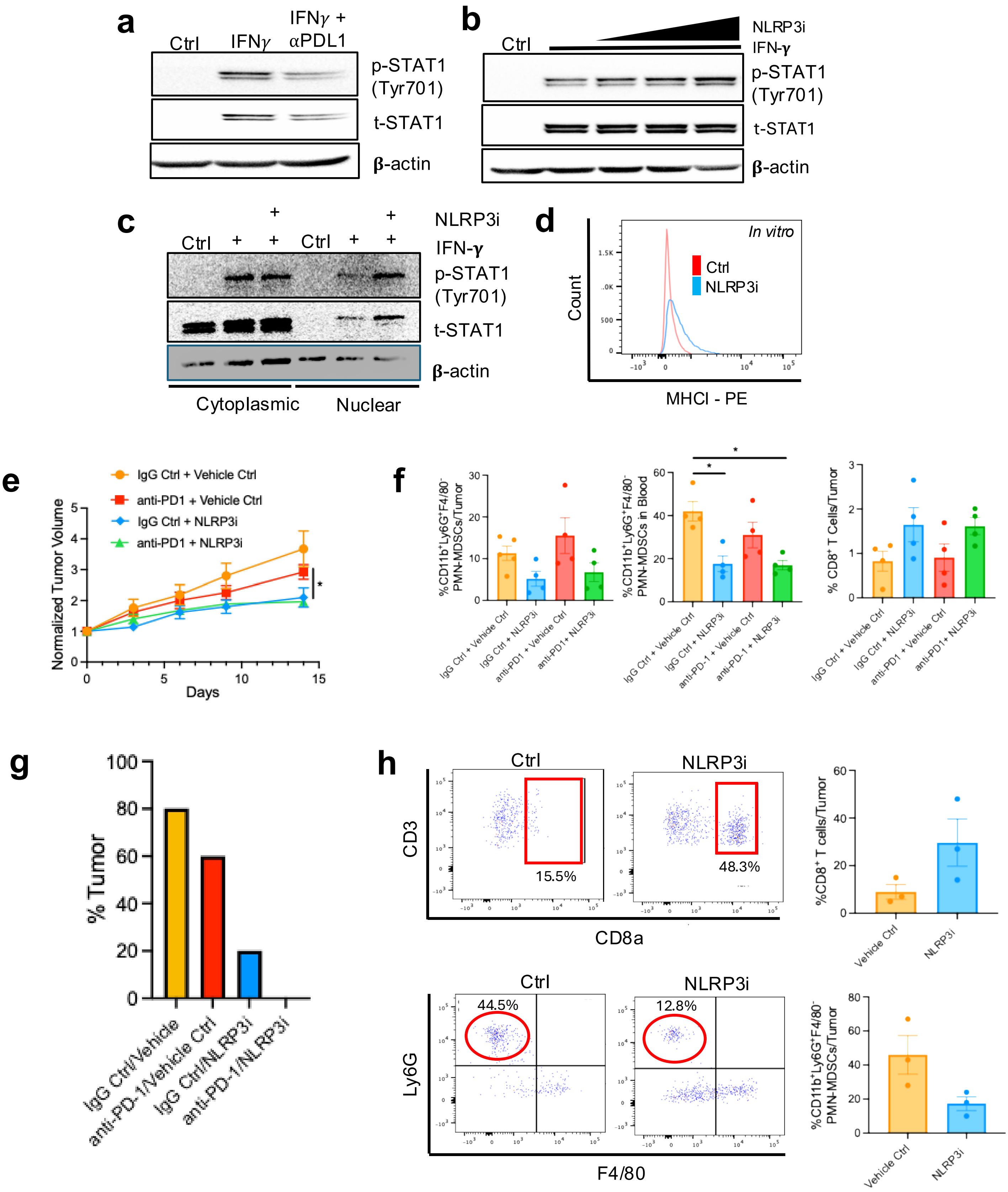
