## Supplementary Tables_BH for "Tumor *NLRP3* Amplification Promotes Immunotherapy Resistance by Suppressing MHC Class I Expression"

### Supplementary Table 1.

| Patient Number | Subject ID | Age (Decade) | Gender | Treatment | Objective Response | PFS (Months) | OS (Months) |
|---|---|---|---|---|---|---|---|
| 1 | H071 | 50s | F | anti-CTLA-4/anti-PD-1 | CR | 60.0 | 60.0 |
| 2 | H042 | 70s | M | anti-PD-1 | CR | 60.0 | 60.0 |
| 3 | V001 | - | - | anti-PD-1 | CR | 60.0 | 60.0 |
| 4 | V002 | - | - | anti-PD-1 | CR | 60.0 | 60.0 |
| 5 | V003 | - | - | anti-CTLA-4 | CR | 60.0 | 60.0 |
| 6 | H097 | 70s | F | anti-CTLA-4/anti-PD-1 | PR | 6.1 | 17.7 |
| 7 | H068 | 50s | M | anti-PD-1 | PR | 60.0 | 60.0 |
| 8 | H031 | 70s | M | anti-PD-1 | PR | 60.0 | 60.0 |
| 9 | H048 | 80s | F | anti-PD-1 | PR | 60.0 | 60.0 |
| 10 | H075 | 70s | M | anti-PD-1 | PR | 60.0 | 60.0 |
| 11 | H081 | 60s | F | anti-CTLA-4/anti-PD-1 | PR | 60.0 | 60.0 |
| 12 | H035 | 60s | M | anti-PD-1 | PR | 60.0 | 60.0 |
| 13 | H103 | 30s | M | anti-CTLA-4/anti-PD-1 | PR | 60.0 | 60.0 |
| 14 | H096 | 70s | M | anti-CTLA-4/anti-PD-1 | PR | 60.0 | 60.0 |
| 15 | H030 | 60s | M | anti-CTLA-4/anti-PD-1 | PR | 60.0 | 60.0 |
| 16 | H055 | 70s | M | anti-PD-1 | PR | 60.0 | 60.0 |
| 17 | H037 | 60s | M | anti-CTLA-4/anti-PD-1 | PR | 60.0 | 60.0 |
| 18 | H053 | 50s | F | anti-PD-1 | PR | 60.0 | 60.0 |
| 19 | H058 | 80s | M | anti-PD-1 | PR | 22.7 | 33.3 |
| 20 | H065 | 50s | M | anti-CTLA-4/anti-PD-1 | PR | 60.0 | 60.0 |
| 21 | H098 | 60s | M | anti-PD-1 | PR | 60.0 | 60.0 |
| 22 | H107 | 60s | F | anti-CTLA-4/anti-PD-1 | PR | 60.0 | 60.0 |
| 23 | H110 | 40s | M | anti-CTLA-4/anti-PD-1 | PR | 11.6 | 60.0 |
| 24 | V012 | - | - | anti-PD-1 | PR | 60.0 | 60.0 |
| 25 | H087 | 20s | F | anti-PD-1 | SD | 2.1 | 29.0 |
| 26 | H052 | 80s | M | anti-PD-1 | SD | 60.0 | 60.0 |
| 27 | V013 | - | - | anti-PD-1 | SD | 14.2 | 52.8 |
| 28 | H109 | 30s | M | anti-CTLA-4/anti-PD-1 | PD | 1.6 | 60.0 |
| 29 | H105 | 60s | F | anti-CTLA-4/anti-PD-1 | PD | 17.0 | 60.0 |
| 30 | H061 | 40s | F | anti-PD-1 | PD | 2.0 | 60.0 |
| 31 | H049 | 70s | M | anti-CTLA-4/anti-PD-1 | PD | 2.7 | 12.3 |
| 32 | H073 | 60s | M | anti-CTLA-4/anti-PD-1 | PD | 3.0 | 13.4 |
| 33 | H045 | 30s | M | anti-PD-1 | PD | 26.8 | 60.0 |
| 34 | H054 | 40s | F | anti-CTLA-4/anti-PD-1 | PD | 0.7 | 23.4 |

#### Supplementary Table 1 (cont).

| Patient Number | Subject ID | Age (Decade) | Gender | Treatment | Objective Response | PFS (Months) | OS (Months) |
|---|---|---|---|---|---|---|---|
| 35 | H059 | 60s | F | anti-PD-1 | SD | 60.0 | 60.0 |
| 36 | H093 | 80s | F | anti-PD-1 | PD | 8.3 | 60.0 |
| 37 | H129 | 60s | F | anti-CTLA-4/anti-PD-1 | SD | 60.0 | 60.0 |
| 38 | V004 | - | - | anti-PD-1 | PD | 7 | 18 |
| 39 | V005 | - | - | anti-PD-1 | PD | 2.8 | 9.8 |
| 40 | V006 | - | - | anti-PD-1 | PD | 2.1 | 2.4 |
| 41 | V007 | - | - | anti-PD-1 | PD | 0.88 | 2.2 |
| 42 | V008 | - | - | anti-PD-1 | PD | 2.7 | 5.7 |
| 43 | V009 | - | - | anti-PD-1 | PD | 2.8 | 9.8 |
| 44 | V010 | - | - | anti-PD-1 | PD | 3.2 | 7.5 |
| 45 | V011 | - | - | anti-CTLA4 | PD | 2.8 | 8.0 |
| 46 | H039 | 70s | M | anti-PD-1 | PD | 2.8 | 60.0 |
| 47 | H032 | 40s | F | anti-PD-1 | PD | 5.4 | 13.2 |
| 48 | H102 | 70s | M | anti-CTLA-4/anti-PD-1 | PD | 2.8 | 60.0 |
| 49 | H043 | 40s | M | anti-PD-1 | PR | 1.1 | 8.0 |
| 50 | H057 | 50s | F | anti-CTLA-4/anti-PD-1 | PR | 11.0 | 60.0 |
| 51 | H076 | 50s | M | anti-PD-1 | PR | 60.0 | 60.0 |
| 52 | H078 | 60s | F | anti-PD-1 | PR | 6.0 | 29.0 |
| 53 | H083 | 60s | F | anti-CTLA-4/anti-PD-1 | PR | 60.0 | 60.0 |
| 54 | H090 | 50s | M | anti-CTLA-4/anti-PD-1 | PR | 60.0 | 60.0 |
| 55 | H104 | 70s | M | anti-PD-1 | PR | 60.0 | 60.0 |
| 56 | H112 | 70s | F | anti-CTLA-4/anti-PD-1 | PR | 60.0 | 60.0 |
| 57 | H113 | 60s | F | anti-PD-1 | PR | 60.0 | 60.0 |
| 58 | H115 | 70s | M | anti-PD-1 | PR | 60.0 | 60.0 |
| 59 | H079 | 80s | M | anti-PD-1 | SD | 10.0 | 16.1 |
| 60 | H066 | 60s | M | anti-PD-1 | SD | 60.0 | 60.0 |
| 61 | H034 | 80s | F | anti-PD-1 | PD | 3.3 | 3.8 |
| 62 | H060 | 60s | F | anti-PD-1 | SD | 5.0 | 60.0 |

### Supplementary Table 2.

| Patient Number | Subject ID | Treatment | RFS (Months) |
|---|---|---|---|
| 1 | H064 | anti-PD-1 | 66.5 |
| 2 | H067 | anti-PD-1 | 65.9 |
| 3 | H074 | anti-PD-1 | 52.0 |
| 4 | H086 | anti-PD-1 | 8.2 |
| 5 | H091 | anti-PD-1 | 56.3 |
| 6 | H092 | anti-PD-1 | 54.9 |
| 7 | H099 | anti-PD-1 | 13.4 |
| 8 | H101 | anti-PD-1 | 7.1 |
| 9 | H106 | anti-PD-1 | 44.1 |
| 10 | H114 | anti-PD-1 | 0.47 |
| 11 | H116 | anti-PD-1 | 27.3 |
| 12 | H117 | anti-PD-1 | 38.5 |
| 13 | H118 | anti-PD-1 | 38.5 |

#### Supplementary Table 3.

| Gene Number | Gene Name |
|---|---|
| 1 | LBP |
| 2 | RASL10B |
| 3 | STRA6 |
| 4 | MATN4 |
| 5 | SERPING1 |
| 6 | KCNMA1 |
| 7 | KCNS1 |
| 8 | NOS2 |
| 9 | HAS1 |
| 10 | C3 |
| 11 | SFRP5 |
| 12 | CEBPD |
| 13 | PDE8B |
| 14 | TMEM108 |
| 15 | HS3ST1 |
| 16 | PDE1A |
| 17 | AEBP1 |
| 18 | CHRDL1 |
| 19 | SULF1 |
| 20 | GPC3 |
| 21 | PTGER3 |
| 22 | SERPINE2 |
| 23 | SRPX2 |
| 24 | KCNK10 |
| 25 | SOD3 |
| 26 | THBS2 |
| 27 | MMP19 |
| 28 | HSD11B1 |
| 29 | NTRK2 |
| 30 | FGF7 |
| 31 | GRIK2 |
| 32 | CHRDL2 |
| 33 | CLEC4D |
| 34 | INHBA |
| 35 | RTN4RL2 |
| 36 | IL1B |
| 37 | APOD |
| 38 | MMP3 |
| 39 | BDKRB1 |
| 40 | CLEC4E |
| 41 | CLSTN2 |
| 42 | LIF |
| 43 | RBPJL |
| 44 | NFASC |
| 45 | CXCL5 |
| 46 | SLC35D3 |
| 47 | SAA3P |
| 48 | IL6 |

**NLRP3-tsig Transcriptional Signature.**

### Supplementary Table 4.

| Patient Number | Subject ID | Age (Decade) | Gender | Tumor Site | PD-L1 CPS | Objective Response | PFS (Months) | OS (Months) |
|---|---|---|---|---|---|---|---|---|
| 1 | 111 | 60s | M | Esophagus | - | PR | 35.6 | 37.3 |
| 2 | 129 | 70s | M | Esophagus | 15 | PR | 13.1 | 24.3 |
| 3 | 114 | 50s | M | Esophagus | 60 | PR | 12.2 | 22.9 |
| 4 | 125 | 30s | M | GE Junction | 0 | SD | 6.6 | 31.4 |
| 5 | 141 | 60s | M | Esophagus | - | PR | 2.1 | 2.1 |
| 6 | 118 | 60s | M | Esophagus | 4 | PD | 2.4 | 2.8 |
| 7 | 127 | 60s | M | Esophagus | 1 | PD | 2.0 | 3.0 |
| 8 | 139 | 60s | M | GE Junction | 20 | PD | 2.3 | 9.2 |
| 9 | 140 | 40s | M | Esophagus | - | PD | 2.3 | 11.6 |
| 10 | 121 | 40s | F | GE Junction | 10 | SD | 4.6 | 15.8 |
| 11 | 120 | 60s | F | GE Junction | 0 | PD | 1.8 | 3.5 |
| 12 | 101 | 60s | M | Esophagus | 40 | CR | 42.1 | 44.6 |
| 13 | 103 | 40s | M | Stomach | 15 | PR | 6.7 | 11.5 |
| 14 | 104 | 50s | M | Esophagus | 3 | PD | 4.2 | 18.4 |
| 15 | 105 | 70s | M | GE Junction | 25 | CR | 28.2 | 28.2 |
| 16 | 106 | 60s | M | Esophagus | 2 | PR | 6.3 | 16.0 |
| 17 | 107 | 50s | M | Esophagus | 1 | PR | 11.2 | 40.9 |
| 18 | 108 | 50s | M | GE Junction | 3 | PR | 8.51 | 8.51 |
| 19 | 109 | 50s | M | Esophagus | 10 | PR | 35.1 | 39.1 |
| 20 | 112 | 50s | M | Esophagus | 0 | CR | 24.5 | 35.3 |
| 21 | 117 | 50s | M | Esophagus | - | PR | 7.6 | 7.6 |
| 22 | 119 | 30s | F | GE Junction | 0 | PR | 9.7 | 30.9 |
| 23 | 122 | 70s | M | Esophagus | 0 | PR | 23.0 | 31.3 |
| 24 | 123 | 40s | M | Stomach | - | PR | 4.8 | 13.9 |
| 25 | 126 | 70s | M | GE Junction | 1 | PR | 14.6 | 14.6 |
| 26 | 128 | 60s | M | Esophagus | - | PR | 8.7 | 29.9 |
| 27 | 130 | 40s | F | Stomach | 5 | SD | 5.8 | 8.5 |
| 28 | 132 | 60s | M | GE Junction | - | PR | 6.6 | 11.6 |
| 29 | 133 | 70s | F | GE Junction | 2 | PD | 2.5 | 2.7 |
| 30 | 134 | 60s | F | Stomach | 0 | SD | 17.5 | 17.5 |
| 31 | 135 | 50s | M | GE Junction | 20 | PD | 2.5 | 4.1 |
| 32 | 136 | 70s | M | GE Junction | 20 | PR | 8.8 | 24.3 |
| 33 | 501 | 70s | M | GE Junction | - | PR | 3.5 | 4.1 |
| 34 | 502 | 60s | M | Stomach | - | PR | 7.1 | 17.7 |
| 35 | 503 | 30s | M | Stomach | - | PR | 12.2 | 25.7 |

### Supplementary Table 5.

| Protein Name | Accession |
|---|---|
| Signal transducer and activator of transcription 1 (STAT1) | sp\|P42225\|STAT1_MOUSE |
| Antigen peptide transporter 2 (TAP2) | sp\|P36371\|TAP2_MOUSE |
| Neurofilament medium polypeptide (NFM) | sp\|P08553\|NFM_MOUSE |
| Interferon-induced transmembrane protein 3 (IFM3) | sp\|Q9CQW9\|IFM3_MOUSE |
| Calsyntenin-1 (CSTN1) | rev_sp\|Q9EPL2\|CSTN1_MOUSE |
| Ankycorbin (RAI14) | sp\|Q9EP71\|RAI14_MOUSE |
| Interferon-induced protein with tetratricopeptide repeats 1 (IFIT1) | sp\|Q64282\|IFIT1_MOUSE |
| Ras-related protein Rab-14 (RAB14) | sp\|Q91V41\|RAB14_MOUSE |
| F-actin-capping protein subunit alpha-2 (CAZA2) | sp\|P47754\|CAZA2_MOUSE |
| Immunity-related GTPase family M protein 1 (IRGM1) | sp\|Q60766\|IRGM1_MOUSE |
| GRB2-associated-binding protein 3 (GAB3) | rev_sp\|Q8BSM5\|GAB3_MOUSE |
| DnaJ homolog subfamily A member 1 (DNJA1) | sp\|P63037\|DNJA1_MOUSE |
| 2'-5'-oligoadenylate synthase 1A (OAS1A) | sp\|P11928\|OAS1A_MOUSE |
| Probable ATP-dependent RNA helicase DDX6 (DDX6) | sp\|P54823\|DDX6_MOUSE |
| Leucine-rich repeat serine/threonine-protein kinase 2 (LRRK2) | sp\|Q5S006\|LRRK2_MOUSE |
| 2'-5'-oligoadenylate synthase-like protein 1 (OASL1) | sp\|Q8VI94\|OASL1_MOUSE |
| Cytochrome c oxidase subunit 7A2, mitochondrial (CX7A2) | sp\|P48771\|CX7A2_MOUSE |
| Ceramide synthase 2 (CERS2) | sp\|Q924Z4\|CERS2_MOUSE |
| Transducin beta-like protein 2 (TBL2) | sp\|Q9R099\|TBL2_MOUSE |
| Torsin-1A-interacting protein 1 (TOIP1) | sp\|Q921T2\|TOIP1_MOUSE |
| Prolow-density lipoprotein receptor-related protein 1 (LRP1) | sp\|Q91ZX7\|LRP1_MOUSE |
| Vigilin (VIGLN) | sp\|Q8VDJ3\|VIGLN_MOUSE |
| T-complex protein 1 subunit beta (TCPB) | sp\|P80314\|TCPB_MOUSE |
| UDP-glucuronosyltransferase 1-6 (UD16) | sp\|Q64435\|UD16_MOUSE |
| G-protein coupled receptor-associated protein LMBRD2 (LMBD2) | sp\|Q8C561\|LMBD2_MOUSE |
| Histone-arginine methyltransferase CARM1 (CARM1) | sp\|Q9WVG6\|CARM1_MOUSE |
| Basement membrane-specific heparan sulfate proteoglycan core protein (PGBM) | sp\|Q05793\|PGBM_MOUSE |

### Supplementary Table 6.

| Gene | Forward | Reverse |
|---|---|---|
| *Stat1* (m) | TCACAGTGGTTCGAGCTTCA | GCAAACGAGACATCATAGGCA |
| *β2m* (m) | ACAGTTCCACCCGCCTCACATT | TAGAAAGACCAGTCCTTGCTGAAG |
| *Nlrc5* (m) | TCAGCCCAGAACAAGTATCC | TGGGCACAGACTTCCATTAG |
| *Actb* (m) | GGCTGTATTCCCCTCCATCG | CCAGTTGGTAACAATGCCATGT |
